# Modeling diagnostic code dropout of schizophrenia in electronic health records improves phenotypic data quality and cross-ancestry transferability of polygenic scores

**DOI:** 10.1101/2025.01.19.25320806

**Authors:** David Burstein, Simone Tomasi, Sanan Venkatesh, VA Million Veteran Program, Mina Rizk, Panos Roussos, Georgios Voloudakis

## Abstract

**Importance:** Researchers commonly use counts of diagnostic codes from EHR-linked biobanks to infer phenotypic status. However, these approaches overlook temporal changes in EHR data, such as the discontinuation or “dropout” of diagnostic codes, which may exacerbate disparities in genomics research, as EHR data quality can be confounded with demographic attributes.

**Objective:** To address this, we propose modeling diagnostic code dropout in EHR data to inform phenotyping for schizophrenia in genomic analyses.

**Design:** We develop and test our diagnostic dropout model by analyzing EHR data from individuals with prior schizophrenia diagnoses. We further validate model performance on a subset of patients whose diagnoses were attained through chart review. Using PRS-CS and existing GWAS summary statistics, we first extrapolate polygenic weights. Then, we apply our dropout model’s outputs to construct a data-driven filter defining our target cohort for measuring polygenic score performance.

**Setting:** Our analysis utilizes EHR and genomic data from the Million Veteran Program.

**Participants:** To model diagnostic dropout in schizophrenia, we leverage data from 12,739 patients with a history of schizophrenia, after excluding outliers. For polygenic score analyses, we incorporate data from a potential pool of 8,385 European ancestry and 6,806 African ancestry patients with a history of schizophrenia.

**Main outcomes and measures:** We compare the performance of our diagnostic dropout model with alternative methodologies both in predicting diagnostic dropout on a holdout set, as well as on chart review labeled data. Using the top differential diagnosis predictors in our model, we select relevant cases by filtering out patients with a prior history of mood or anxiety disorders. We then test the impact of applying different filters for measuring polygenic score performance.

**Results:** When evaluated on chart review-labeled data, our model improves the area under the precision-recall curve (AUPRC) by 9.6% compared to competing methods. By applying our data-driven filter for schizophrenia, we achieve a 62% increase in the association effect size when transferring a European polygenic score to an African ancestry target cohort.

**Conclusions and Relevance:** These findings highlight the potential of modeling diagnostic code dropout to enhance the phenotypic quality of EHR-linked biobank data, advancing more equitable and accurate genomics research across diverse populations.

**Key Points:** *Question:* Can we leverage temporal changes in electronic health record (EHR) data to improve schizophrenia case selection for genomic studies?

*Findings:* We trained an XGBoost model on EHR data from 12,739 patients to predict schizophrenia diagnostic code dropout in the Million Veteran Program. By excluding cases with conditions associated with diagnostic dropout, we achieved a 62% increase in effect size when applying polygenic weights to an African ancestry target cohort. Filtering based on substance use, a common approach, yielded minimal gains.

*Meaning:* Modeling diagnostic code dropout enhances the phenotypic quality of EHR-linked biobank data, and promotes equitable genomics research across diverse populations.

## Introduction

While the lack of participant diversity in genome-wide association studies diminishes the equitability of leveraging polygenic scores for informing diagnoses^1^, large electronic health record (EHR) biobanks hold great promise for promoting diversity in human genetics studies^2^. Nevertheless, extracting phenotypic definitions from EHR data presents serious challenges as EHR data quality can vary in completeness across different patients^3^. Furthermore, cryptically embedded biases in EHRs can compromise data quality and exacerbate disparities in genetics research. For example, research has demonstrated that Black individuals are more likely to receive a schizophrenia diagnosis than their white counterparts, suggesting that data quality can vary across different demographic groups^4–6^. As a result, noisy data can diminish or bias the genetic signal in both genome-wide association and polygenic score performance for patients with diverse backgrounds^7^. Consequently, we need methodologies that can robustly identify cases from EHRs to mitigate the impact of these hidden disparities in our genetic analyses.

Arguably the most popular methodology for assigning cases from EHR data leverages diagnosis code counts to identify cases, where researchers define cases by requiring 2 or more related diagnosis codes to appear in a patient’s chart^8,9^. Alternative methodologies, such as multimodal automated phenotyping (MAP^10^), use mixture models, integrating both structured (e.g., diagnostic code counts) and semistructured (e.g., codes from medical narrative) data to estimate the probability of a patient having the diagnosis and if needed the diagnostic threshold value. However, these approaches do not account for competing diagnoses that could interfere with a clinician’s ability to correctly diagnose a patient with schizophrenia.

Here, we propose a diagnostic dropout modeling framework to improve the equitability of genomics research by identifying data quality issues that affect patients across different demographic backgrounds. More specifically, we train an XGBoost (Extreme Gradient Boosting) model to predict the discontinuation or “dropout” of diagnostic codes for schizophrenia using prior medication and diagnoses from the EHR data in the Million Veteran Program. We benchmark our methodology against both MAP and diagnosis code count methodologies using two holdout samples, one for predicting the presence of future schizophrenia diagnosis codes and the other with diagnoses determined through chart review. Finally, we investigate how improving phenotyping definitions for schizophrenia impacts the transferability of polygenic risk scores by applying a data-driven filter for schizophrenia based on the top risk factors for diagnostic dropout in our model.

## Methods

### Study Participants

This study was approved by the Veterans Affairs (VA) Central Institutional Review Board, and all patients provided written informed consent. We refer the reader to prior work describing the cohort in further detail^6,11,12^.

### MVP

The Million Veteran Program (MVP) cohort has been previously described^6,11–14^. Analysis of MVP data uses the latest EHR tables, v23.1, covering 33,699 individuals with schizophrenia diagnosis codes up to September 2023. After filtering for diagnosis frequency, we leverage 12,739 individuals for training and testing our phenotypic model, with records covering a median of 21.6 years (interquartile range, IQR = 7.62), a median 776 outpatient visits (IQR = 823) and a median density of 38.7 outpatient visits per year (IQR = 40.32). This cohort includes 146 individuals with chart review labels. For genetic analyses, we utilize release 4 of the data, consisting of 8,385 European and 6,806 African ancestry patients with 2 or more schizophrenia diagnosis codes (**Supplementary Table S1**). Genotyping, quality control and imputation in MVP is handled by a dedicated data team. For further details, see the **eMethods**.

### CSP #572

The CSP #572 cohort has been previously described^6^. Briefly, participants were recruited through participating VA hospitals between 2011 and 2020. Structural Clinical Interviews for the DSM were performed for all patients with schizophrenia. For the scope of this work, we leverage African ancestry genome-wide summary statistics using cases from CSP #572 for schizophrenia to construct a polygenic score.

### Modeling Diagnostic Code Dropout

To remedy data quality issues for resolving diagnostic inaccuracies in this EHR dataset, we leverage XGBoost^15^, a machine learning methodology suited for capturing complex non-linear relationships between features while combining boosting with regularization, to predict future schizophrenia diagnosis codes based on prior prescriptions and diagnosis codes (**Figure 1**). True cases of schizophrenia and other chronic diagnoses are expected to continue to receive schizophrenia diagnoses and prescriptions throughout their life. We separate counts of diagnoses and prescriptions based on whether they occurred more than 10 years before the most recent encounter. “Future schizophrenia diagnosis codes” are defined as at least 2 schizophrenia codes in the five most recent years in the EHR, using the phecode definition^16^. For training our diagnostic dropout model, we utilize individuals with MAP scores and at least one schizophrenia diagnosis code prior to the 5 most recent years of the patient record (n=12,739). (**eMethods)**.

**Figure 1:**
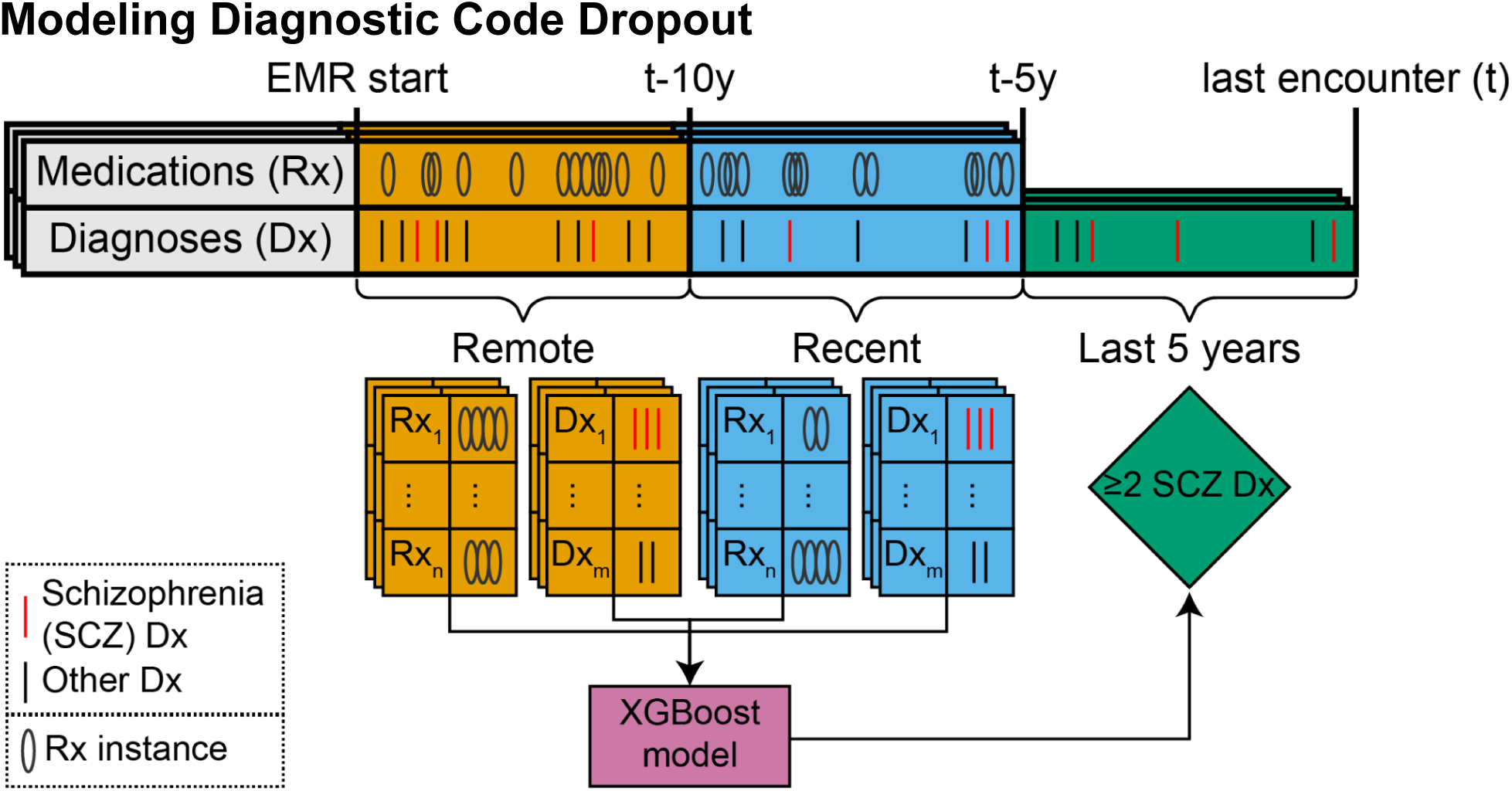
Schematic of model. A schematic diagram illustrating how we define diagnostic dropout for schizophrenia, where we expect true schizophrenia cases to continue to have schizophrenia diagnoses based on information within the last 5 years since the most recent outpatient encounter in the patient’s electronic health record, using the 2+ diagnosis code count rule. To train our predictive model, we partition the patient’s electronic health record in time from the most recent outpatient encounter. The color indicates the timeframe for defining diagnosis and prescription counts. Directional arrows indicate that prior diagnosis and prescription data are used to predict recent schizophrenia counts. Abbreviations: Rx: prescriptions; Dx: diagnoses; SCZ: schizophrenia; EMR: electronic medical records; 5y: 5 years; 10y: 10 years.

### Chart Review Data Validation

We validate the model’s predictions by comparing them against manually verified EHR information using chart review. We define a subset of 146 charts, randomly partitioned and assigned to two expert psychiatrists. (**Supplementary Table S2-S3, Supplementary Figure S1, eMethods)**. We define a case if a patient received “Yes schizophrenia” ratings from both psychiatrists. We also train a logistic regression model to predict “Yes schizophrenia” ratings, leveraging prior diagnosis code counts over a five-year period for schizophrenia, substance use disorders, anxiety disorders, mood disorders and adjustment disorders.

### Assessment of Data-driven Phenotypes with Polygenic Scores

To test our hypothesis that robustly defined phenotypes will enhance the polygenic signal, we construct a data-driven filter by excluding schizophrenia cases with top negatively associated diagnoses from the diagnostic dropout model. We consider different case definitions based on MAP and diagnosis code count thresholds for African and European ancestry validation cohorts from the Million Veteran Program. We employ PRS-CS^17^ to construct polygenic scores from European ancestry GWAS summary statistics from the PGC^18^ as well as African ancestry GWAS summary statistics from CSP 572^19^. We quantify polygenic score performance using logistic regression adjusting for sex, age and top ten ancestry principal components (**eMethods**).

### Assessment of Data-driven Phenotypes by Measuring Dilution (PheMED)

We leverage PheMED to measure how data-driven filters can impact the distribution of effect sizes on genome-wide association summary statistics^7^. More specifically, integrating noisier data with inaccurately labeled cases can dilute effect sizes by the same factor across all variants correlated with the phenotype. Unlike polygenic score validation analyses, which include significant measurement error by retaining uncorrelated SNPs, PheMED can infer dilution estimates without direct knowledge of the causal or correlated SNPs in a GWAS. For further details, on implementation see **eMethods**.

## Results

### Predicting Diagnostic Dropout in Electronic Health Record Data

In **Supplementary Figure S2 and Table S4,** we examine the top predictors of our XGBoost Model for predicting future schizophrenia diagnosis codes. Unsurprisingly, our model identifies prior schizophrenia diagnoses as the most important feature in predicting future schizophrenia diagnosis codes, with recent counts being more informative, while schizophrenia MAP scores capture supplemental information from medical narratives (**Supplementary Figure S2a, S2d**). Additionally, our model also leverages differential diagnoses like mood disorders^20,21^ and anxiety disorders^22^ (**Supplementary Figures S2b)** to inform predictions of diagnostic dropout for schizophrenia. These differential diagnoses are negatively associated with future schizophrenia diagnoses as indicated in the accumulated local effect (ALE) plots (**Supplementary Figure S2b-d**). Our model also identifies several medications, including second-generation antipsychotics such as olanzapine, known for its strong efficacy in managing schizophrenia symptoms. For further details on the medications selected by our model, see the **Supplementary Results.**

### EHR and Chart Review Dropout Model Validation

To assess the performance of our diagnostic dropout model, we measure model performance for predicting future schizophrenia diagnosis codes on a holdout set comprising 10% of the cohort. We utilize MAP as the primary benchmark for evaluating model performance, as MAP incorporates both diagnosis counts and concept unique identifiers from patient notes into their predictions. We observe a 3.4% absolute increase in AUROC and a 3% increase in AUPRC when selecting the diagnostic dropout model over MAP (**Supplementary Figure S3**). Unsurprisingly, we observe larger increases in AUROC and AUPRC of 5.0% and 4.1% respectively when comparing our methodology to a classifier that ranks potential cases by diagnosis code counts (**Supplementary Figure S4**). We perform logistic regression to predict future schizophrenia codes on the holdout set and observe a statistically significant association with our diagnostic dropout model scores (**β** = 1.128, *p* = 6.71 × 10^−30^), even when adjusting for MAP scores and log transformed diagnosis code counts.

As diagnostic dropout is a proxy measurement for schizophrenia misdiagnosis, we then measure our model performance on a separate holdout set of 146 individuals (**Supplementary Table S2-S3**; **Supplementary Figure S5, Methods**) where diagnoses were attained through chart review. Chart review labels were consistent across reviewers. When we simplified the classification to two options (yes or no), Cohen’s interrater kappa (κ), equaled 0.837, reflecting a high level of agreement, consistent with prior literature ^23,24^ (**eMethods)**. Here, we observe more substantial differences in model performance. We attain a 5.7% absolute increase in AUROC and a 9.6% increase in AUPRC when selecting the diagnostic dropout model over MAP for predicting diagnostic status attained through chart review (**Figure 2**). When comparing our diagnostic dropout model with diagnosis counts, we observe larger absolute increases of 11.8% for AUROC and 10.7% for AUPRC, respectively (**Supplementary Figure S6**). We also perform logistic regression to predict chart review labels and observe a statistically significant association with our diagnostic dropout model probabilities (**β** = .601, *p* = .001), even when adjusting for MAP scores and log transformed diagnosis code counts.

**Figure 2:**
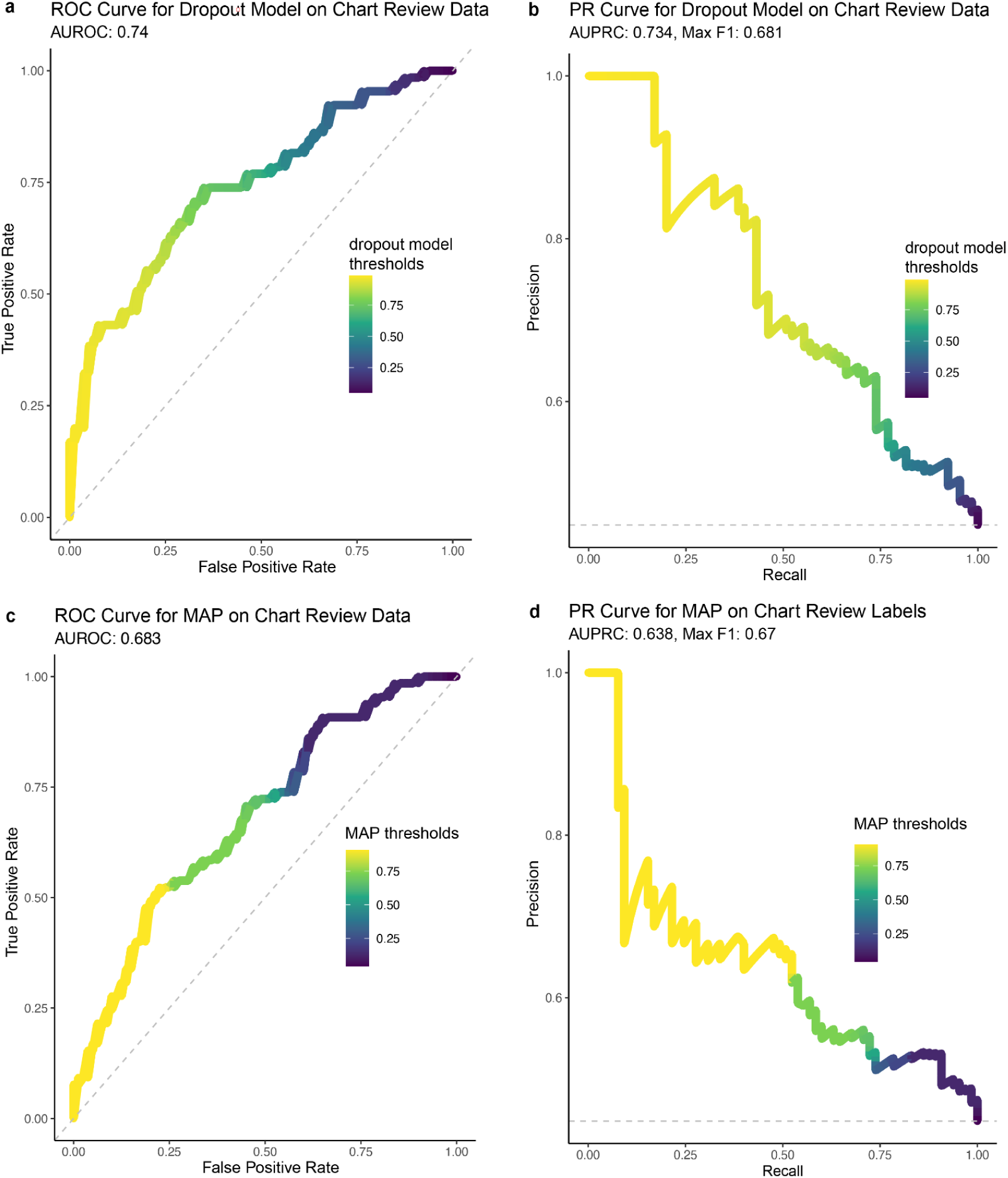
Performance Predicting Chart Review Labels. Receiver operator (ROC) and precision recall (PR) curves assessing the performance of our diagnostic dropout model **(a-b)** and MAP model **(c-d)** on predicting diagnoses attained through chart review. For the precision-recall curves **(b,d)**, the x-axis identifies the recall value, and the y-axis identifies the precision. A dashed line highlights the baseline precision of a random classifier. In the receiver operator curves, the x-axis identifies the false positive rate and the y-axis indicates the true positive rate. The dashed diagonal line corresponds to the performance of a random classifier. Points are color-coded identifying the threshold that yields the corresponding precision-recall or false positive rate/true positive rate value.

### Leveraging the Dropout Model for Schizophrenia Case Selection in Genetic Studies

Even though our diagnostic dropout model yields substantial improvements compared to the MAP and phecode count benchmarks, we observe that our model overemphasizes prior schizophrenia diagnosis code counts and underemphasizes relevant differential diagnoses (**Supplementary Figure S2**), which will likely produce noisy definitions for schizophrenia in genomic analyses. Since our model was designed to predict future diagnosis code counts and not true schizophrenia status, we instead focus on constructing a data-driven filter to flag potentially noisy data, as even modestly noisy phenotypes can dilute the signal when performing polygenic score (PGS) validation^7^. Consequently, we construct an auxiliary logistic regression model to measure statistical significance of the top differential diagnoses in the model. We find that only “mood disorders”, which encompasses both unipolar and bipolar depression, and “anxiety disorders” achieve statistical significance after accounting for multiple test correction, (anxiety disorders *p* = 1.52×10^−26^; mood disorders *p* = 2.49×10^−46^) (**Supplementary Figure S7)**. Furthermore, prior mood disorders and anxiety disorders reached or nearly reached nominal significance for predicting chart-reviewed schizophrenia cases (anxiety disorders *p =* .045, mood disorders *p* = .061, one-sided) (**Supplementary Table S4, Supplementary Figure S8, Methods)** Based on prior literature^25^, we also tested if there was an association between substance use disorders and schizophrenia chart review scores or diagnostic dropout in the electronic health record data, but we did not achieve statistical significance in these analyses. For subsequent analyses, we define our data-driven definition for schizophrenia to construct our target cohorts for aggregate polygenic risk stratification, where we exclude cases that have codes for the statistically significant differential diagnoses associated with diagnostic dropout.

### Polygenic Score Performance of Diagnostic Dropout Filter

We observe significant improvements in the PGS-trait association effect sizes when the base (GWAS used for PGS estimation) and target (individuals for which the PGS is estimated) cohorts share the same ancestry for both European (EUR) and African (AFR) ancestries on MAP-defined schizophrenia cases (**Figures 3a, Supplemental Figure S9, Supplementary Table S5, Methods**). For example, our PGS log odds ratio increases from .599 to .726 when the training and target cohort both share European ancestry *(p* = 4.59 ×10^−6^, 21.2% relative increase). Furthermore, applying our diagnostic dropout filter improves PGS transferability, yielding the largest relative gains when validating EUR GWAS-derived PGS on an AFR target cohort (**β** increases from .120 to.195, 62.2% relative increase, *p* = 7.55 ×10^−3^ with MAP defined cases). In contrast, we observe no significant improvement when validating AFR GWAS-derived PGS on a EUR target cohort. We also observe similar improvements when utilizing a 2+ diagnosis code count rule for defining schizophrenia (**Figure 3b, Supplemental Figure S9, Supplementary Table S5**), where our PGS log odds ratio increases from .526 to .673 *(p* = 6.86 ×10^−8^, 27.95% relative increase) when the training and target cohort both share European ancestry and from .114 to .190 *(p* = 7.86 ×10^−3^, 67.66% relative increase) when transferring European ancestry scores to an African ancestry cohort. PheMED^7^ analysis demonstrates that genome-wide summary statistics from cases not passing the diagnostic dropout filter had, on average, SNP-phenotype association effect sizes 1.328 times smaller (*p* =1.84 ×10^−4^) than those from cases that passed the filter (**Supplemental Table S6**).

**Figure 3:**
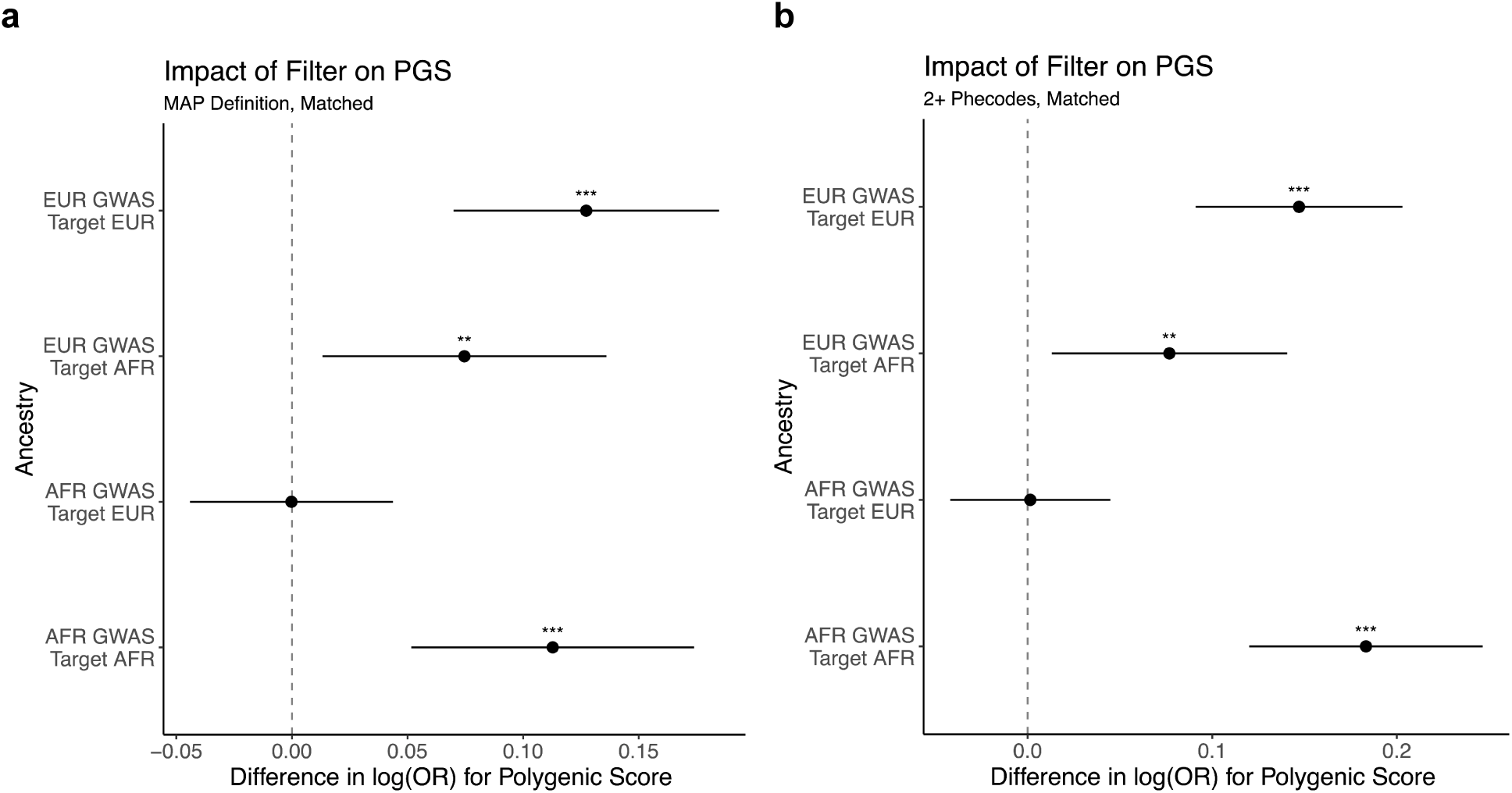
Improvement of PGS Performance when applying Data-Driven Filter. **(a-b)** Forest plots for the differences in effect size of polygenic score predictor between applying and not applying our data-driven filter for schizophrenia cases based on the diagnostic dropout model. The x-axis indicates the difference in the log odds ratio of the polygenic score. The y-axis corresponds to the training cohort GWAS for constructing the PGS as well as the target cohort. While we plot two-sided confidence intervals, statistical tests are one-sided. A dashed vertical line corresponds to the null hypothesis of no difference in effect between filters. P-values are annotated as follows: * p < .05, ** p < .01, *** p < .001. Plot (**a**) uses the MAP definition to define schizophrenia cases, whereas plot (**b**) uses the diagnosis code count to define schizophrenia cases. Cases were matched to mitigate differences in distributions of MAP scores or diagnosis code counts between cases that passed and failed the filter (**Methods**).

### Robustness of Results across Different Thresholds

We assess our diagnostic dropout filter across varying diagnostic stringency thresholds, (as selecting larger MAP probabilities should increase the certainty that a patient has schizophrenia). Surprisingly, PGS performance increases from our filter are robust across all tested thresholds (**Figures 4a-4b**). We observe these improvements both when the target cohort’s genetic ancestry matched the training cohort and when transferring EUR GWAS-derived PGS to an AFR target cohort. We do not however observe a statistically significant improvement for AFR GWAS-derived PGS applied to a EUR target cohort, similar to our initial analysis.

**Figure 4:**
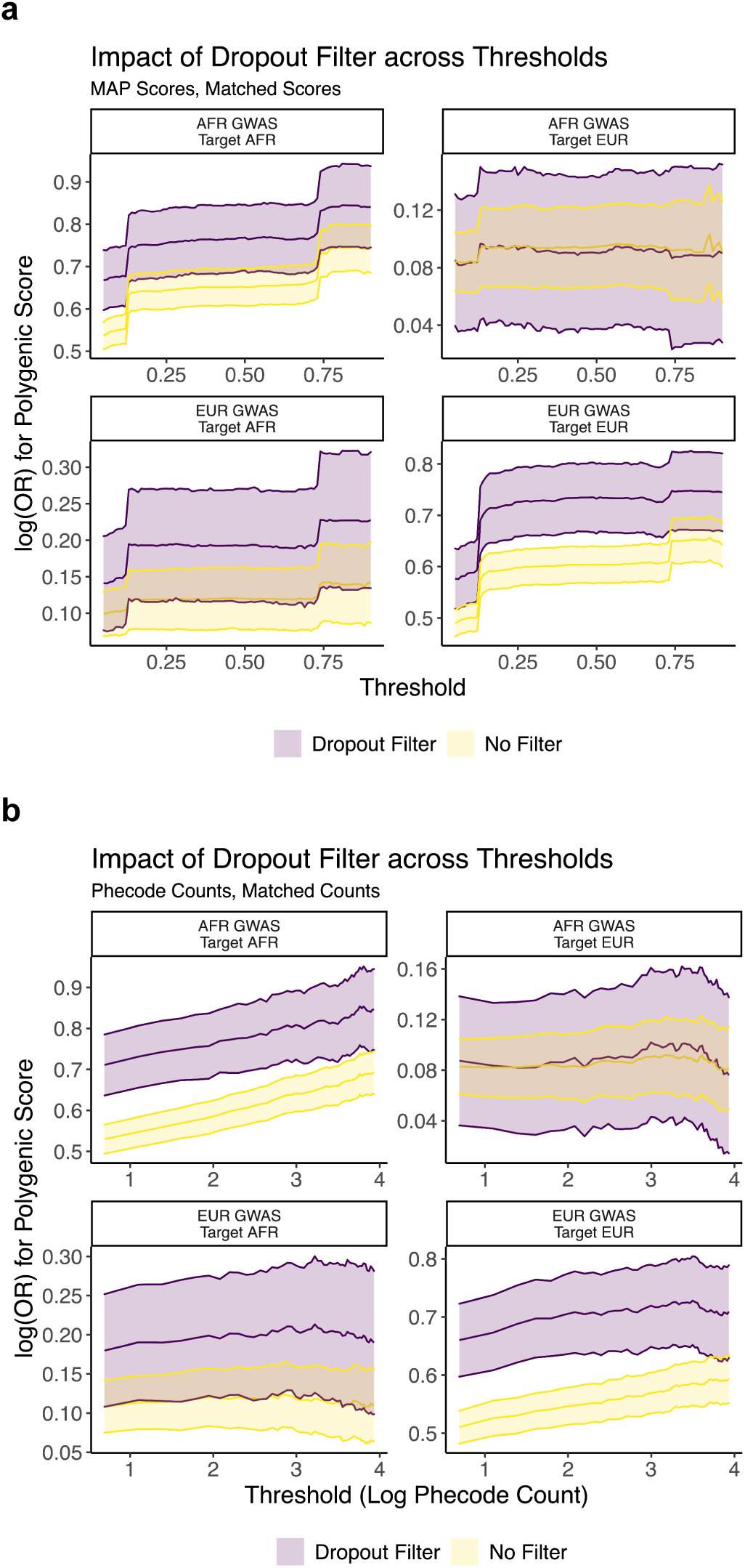
Improvement of PGS Performance across Thresholds. **(a-b)** Here, we plot the effect size of the polygenic risk score (y-axis) across a range of thresholds (x-axis) for defining cases based on the MAP definition (**a**) or the phecode (grouped diagnosis (dx) code) count definition (**b**). The color of the curve corresponds to whether the data driven filter was applied for filtering cases. We also provide 95% confidence intervals for the curves in the plot as well. The subtitle of the plot indicates the training cohort GWAS for constructing the PGS as well as the target cohort.

### Comparison Against Existing Filtering Strategies

We compare our data-driven filtering approach to other proposed filters, including a strict filter for bipolar disorder filter (a highly co-heritable disorder^26–29^) and the mode 5 filter, which uses the prevailing diagnosis of either schizophrenia or bipolar disorder from the last 5 encounters (**Figure 5**). Our filter outperforms these methods. When applying European ancestry-derived PGS on MAP-defined cases with European ancestry, our data-driven filter increases the log odds ratio by .093 (*p =* 7.15 ×10^−4^) and .125 (*p* = 4.79 ×10^−6^) when compared to a strict bipolar disorder and a mode 5 filter for bipolar disorder respectively. Furthermore, when transferring polygenic scores from a European ancestry training cohort to an African ancestry target cohort, our data-driven filter increases the log odds ratio by .05 (*p =* .0483) compared to the strict bipolar disorder filter and by .0780 (*p* = 8.27 ×10^−3^), compared to the mode 5 bipolar disorder filter.

**Figure 5:**
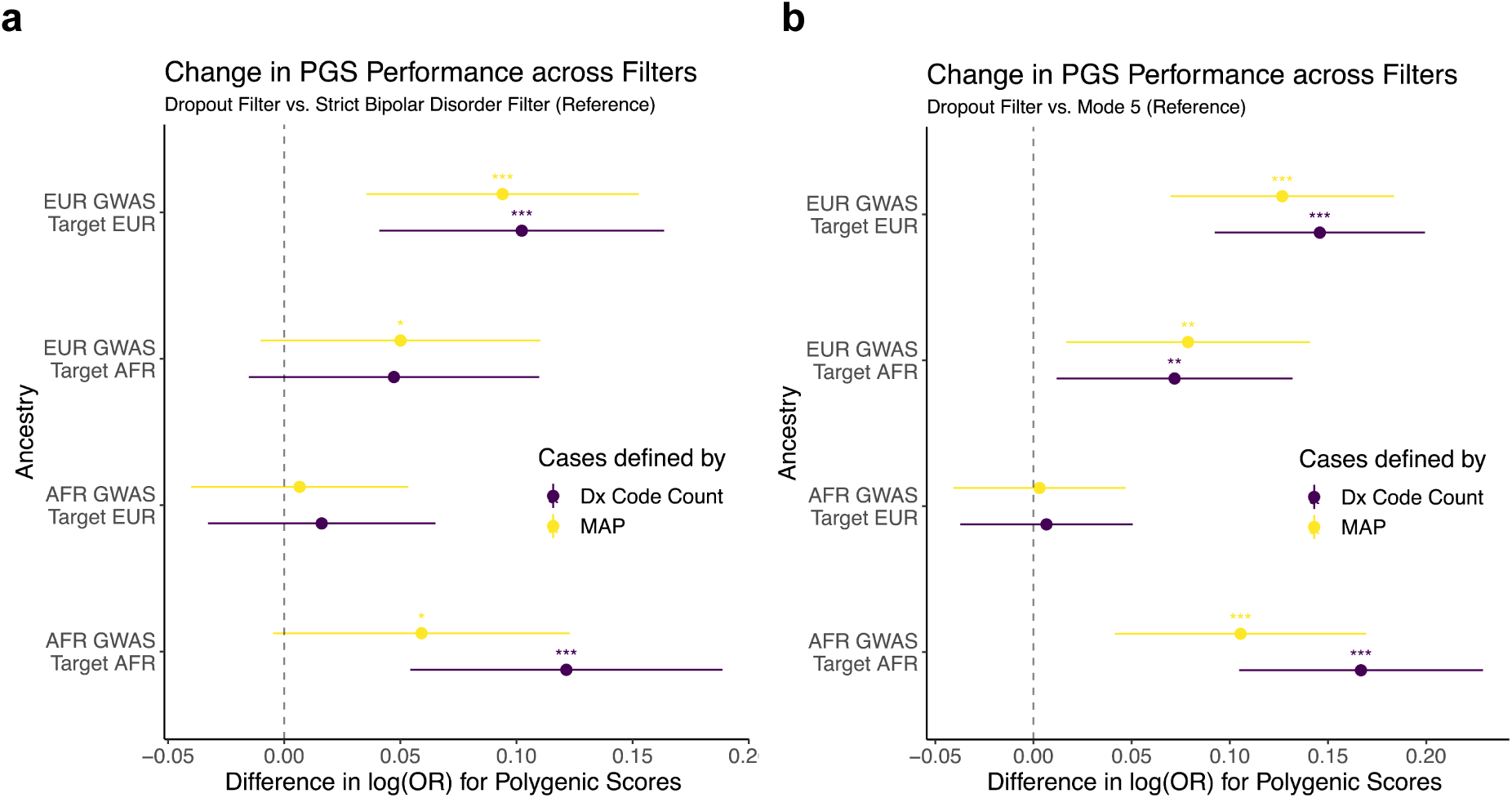
Comparing Data-Driven Filtering approaches. **(a-b)** Forest plots for the differences in effect size of polygenic score predictor among applying different filters for schizophrenia cases. In **(a)** we compare the dropout filter against a strict bipolar disorder filter and in **(b)** we compare the diagnostic dropout filter against the Mode 5 filter, removing cases where bipolar disorder diagnoses is more commonly observed than schizophrenia diagnoses across the five most recent bipolar/schizophrenia diagnoses. The x-axis indicates the difference in the log odds ratio of the polygenic score. The y-axis corresponds to the training cohort GWAS for constructing the PGS as well as the target cohort. The color of the confidence interval corresponds to whether the MAP or diagnosis code (phecode) count definition was used for identifying schizophrenia cases. While we plot two-sided confidence intervals, statistical tests are one-sided. A dashed vertical line corresponds to the null hypothesis of no difference in effect between filters. P-values are annotated as follows: * p < .05, ** p < .01, *** p < .001.

We also explored the impact of a substance use filter on top of a bipolar disorder filter^25^. We find that applying a substance use filter often resulted in weaker effect sizes (**Supplementary Figures S11-S12, Supplementary Table S5**). For instance, the substance use filter *decreased* the PGS effect size by 13.9% (*p =* .0124) compared to a stringent bipolar disorder baseline filter, when the training and target cohorts both shared African ancestry on MAP-defined cases. This aligns with our finding that substance use is positively correlated with future schizophrenia diagnoses and may not be suitable for exclusion in genomic analyses (**Supplementary Figure S7, Supplementary Table S5)**. These findings underscore the importance of data-driven filters to enhance polygenic score analyses.

## Discussion

To mitigate disparities in data quality across demographic groups, we present a novel approach for improving phenotypic data quality by using machine learning to predict diagnostic dropout for schizophrenia in MVP. This model identifies top confounding diagnoses contributing to diagnostic dropout and low chart review ratings for schizophrenia. From these differential diagnoses, we construct a data-driven filter to improve phenotypic data quality. We observe a 62.2% increase in the PGS association effect size when transferring European ancestry-derived scores to an African ancestry validation cohort and a 17.4% and 21.2% increase in effect size for African and European ancestries respectively when the target cohort shares the same ancestry as the training cohort. Our results suggest that to enhance PGS-based risk stratification for schizophrenia, both within and across ancestries, we must use approaches that produce a “cleaner” phenotype for PGS estimation. This incentive conflicts with two major priorities in traditional GWAS studies: maximizing sample size for locus discovery power and using simple phenotypic definitions across cohorts by large consortia. At this juncture, we discuss the broader implications of our work.

Our schizophrenia diagnostic dropout model demonstrates that differential diagnoses such as mood disorders, and anxiety disorders increase the probability of diagnostic dropout for schizophrenia. Notably, a prior study found that up to 40% of schizophrenia patients will also meet the diagnostic criteria for major depression within their lifetime^30,31^. Consequently, while researchers may be inclined to retain patients with both diagnoses, we recommend caution against overly inclusive trait definitions, as noisy phenotypes can dilute findings in GWAS and PGS analyses^7^. Our work highlights the potential consequences when researchers do not apply robust phenotyping approaches for assessing PGS performance. By leveraging data-driven techniques to select 35% of the potentially relevant schizophrenia cases (**Supplementary Table S1**), we achieved a 62% relative increase in the effect size when transferring findings from a European ancestry study to an African ancestry cohort.

Subsequently, we compare the efficacy of our data-driven filter with other approaches, including a strict bipolar disorder exclusion and a mode-based method (Mode 5)^6^. While Mode 5 retains more cases, it’s prone to noise due to a small sample of recent diagnoses. Our filter outperforms these, increasing PGS effect sizes by 15.4% to 20.9% in European ancestry cohorts and 35.4% to 65.6% in African ancestry cohorts. Consequently, our data-driven approach for identifying diagnoses that contribute to diagnostic dropout for schizophrenia is crucial for mitigating disparities in research, where healthcare disparities impact data quality in genomic analyses.

Prior research has advocated for substance use filters due to potential misdiagnosis with schizophrenia, as substance use can induce psychosis^25,32^. However, such a filter could promote inequities in genomic research, as Blacks in the United States, are more likely to be overdiagnosed with substance use disorders^19,33,34^. Substance use disorders are highly comorbid with schizophrenia and a history of substance use could support a schizophrenia diagnosis^35^. Our analyses show positive associations between a history of substance use diagnoses and future schizophrenia codes in a patient’s chart. This trend aligns with our chart review analysis, where substance use disorder codes were positively (although not statistically significantly) correlated with higher chart review ratings **(Supplementary Figures S7-S8)**. The lack of evidence between schizophrenia misdiagnosis and substance use disorders suggests that such a filter would unnecessarily exclude 47% of schizophrenia patients with concomitant substance use disorders^35^. Applying this filter in PGS analyses shows either statistically significant decreases or no evidence of a change in the effect size for nearly all training-target cohort combinations. This finding aligns with our diagnostic dropout model, where substance use was not a top feature in predicting diagnostic dropout. As a result, our data-driven approach for modeling diagnostic dropout provides researchers with a valuable tool for identifying relevant filters that can enhance polygenic signals across patients with diverse backgrounds.

### Limitations

Notably, diagnostic dropout models work best for chronic diagnoses, like schizophrenia, where patients with the diagnosis receive lifelong support to help mitigate the severity of both their persistent and recurrent symptoms. Furthermore, while our diagnostic model was trained using data from MAP, we stress that our modeling framework does not strictly require MAP probabilities if concept unique identifier counts from patient notes are not readily available to researchers. When comparing findings across different EHR-linked biobanks, results may differ. For example, whereas anxiety disorders served as a top predictor for diagnostic dropout in our model, anxiety disorders were not commonly found as a common misdiagnosis in an electronic health record study from a cohort in Spain^20^. Demographically, our cohort is distinct from other biobanks, as it is predominantly composed of male participants with either European or African ancestry. Finally, negative mental health outcomes, including depression and anxiety which are part of the data-driven filter, are also positively associated with self-reported stressors and racism^36,37^. Consequently, it is feasible that a significant percentage of the increased PGS performance is not attributable to the use of a “cleaner” phenotype, but rather from studying individuals under less stress.

## Conclusions

Achieving greater certainty in case status in EHRs is crucial for mitigating disparities in EHR-linked biobanks. Modeling diagnostic dropout probabilities improves phenotypic data quality, generating a 9.6% and 10.7% absolute increase in AUPRC compared to MAP and phecode counts, respectively. Furthermore, we observe a 62% relative increase in polygenic score effect sizes when transferring European ancestry weights to an African ancestry target cohort. Our findings are robust across MAP and PheCode definitions. As a result, we anticipate that modeling diagnostic dropout will serve as a valuable technique for researchers to mitigate cryptically embedded biases in genomic analysis from EHR-linked biobanks.

## Data Availability

GWAS summary statistics and polygenic score weights generated from this paper will be shared through dbGAP upon publication.

## Acknowledgements

This research is based on data from the Million Veteran Program, Office of Research and Development, Veterans Health Administration, and was supported by awards #MVP076 (PR) and MVP#096 (DB). This publication does not represent the views of the Department of Veteran Affairs or the United States Government. This study was also supported by the National Institutes of Health (NIH), Bethesda, MD under award numbers: K08MH122911 (GV), R01AG078657 (GV), R01AG067025 (PR), R01AG082185 (PR), R01AG065582 (PR), R01AG067025 (PR), R01MH125246 (PR) and by the Veterans Affairs Merit grants BX006500 (DB) and BX004189 (PR). We would also like to thank Lauren Costa, Rahul Sangar and Anne Yuk-Lam Ho of the MVP Phenomics Data Core for their support in establishing the chart review pipeline, as well as Tim Bigdeli and Phil Harvey for providing the summary statistics from CSP #572.

