## Supplementary Figures, Methods for "Modeling diagnostic code dropout of schizophrenia in electronic health records improves phenotypic data quality and cross-ancestry transferability of polygenic scores"

### Supplementary Information

#### Table of Contents for Supplementary Information


**Table of Contents for Supplementary Information** **1**
**Supplementary Figures** **2**
Figure S1 2
Figure S2 3
Figure S3 4
Figure S4 5
Figure S5 6
Figure S6 7
Figure S7 8
Figure S8 9
Figure S9 10
Figure S10 11
Figure S11 12
Figure S12 13
**eMethods** **14**
CSP 572 and MVP 14
Predicting Diagnostic Dropout 14
Chart Review Validation 14
Genotyping, quality control and imputation 15
Assessment of Data-driven Phenotypes with Polygenic Scores 15
Measuring Dilution with PheMED 16
**Supplementary Results** **16**
Top Medications in Model 16
Rater Agreement 17
Selecting Top Differential Diagnoses 17
**Supplementary Acknowledgements** **18**
**Supplementary References** **20**

### Supplementary Figures

#### Figure S1

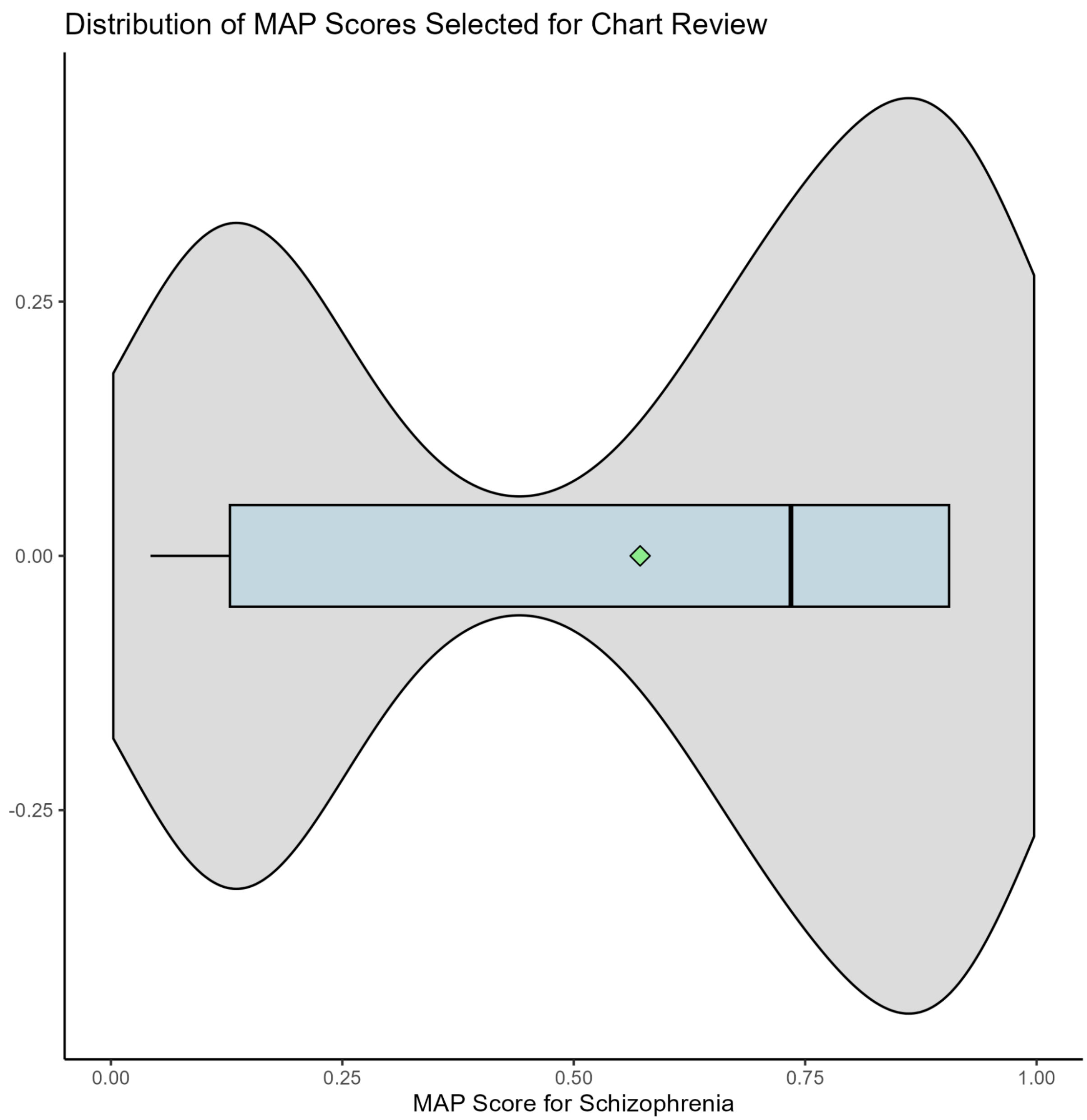


**Figure S1. Distribution of MAP scores for chart review.**
Visualizing the distribution of MAP scores for chart review with a violin plot in gray as well as a boxplot in blue. The green diamond indicates the mean of the distribution. The x-axis identifies the corresponding MAP score.

### Figure S2

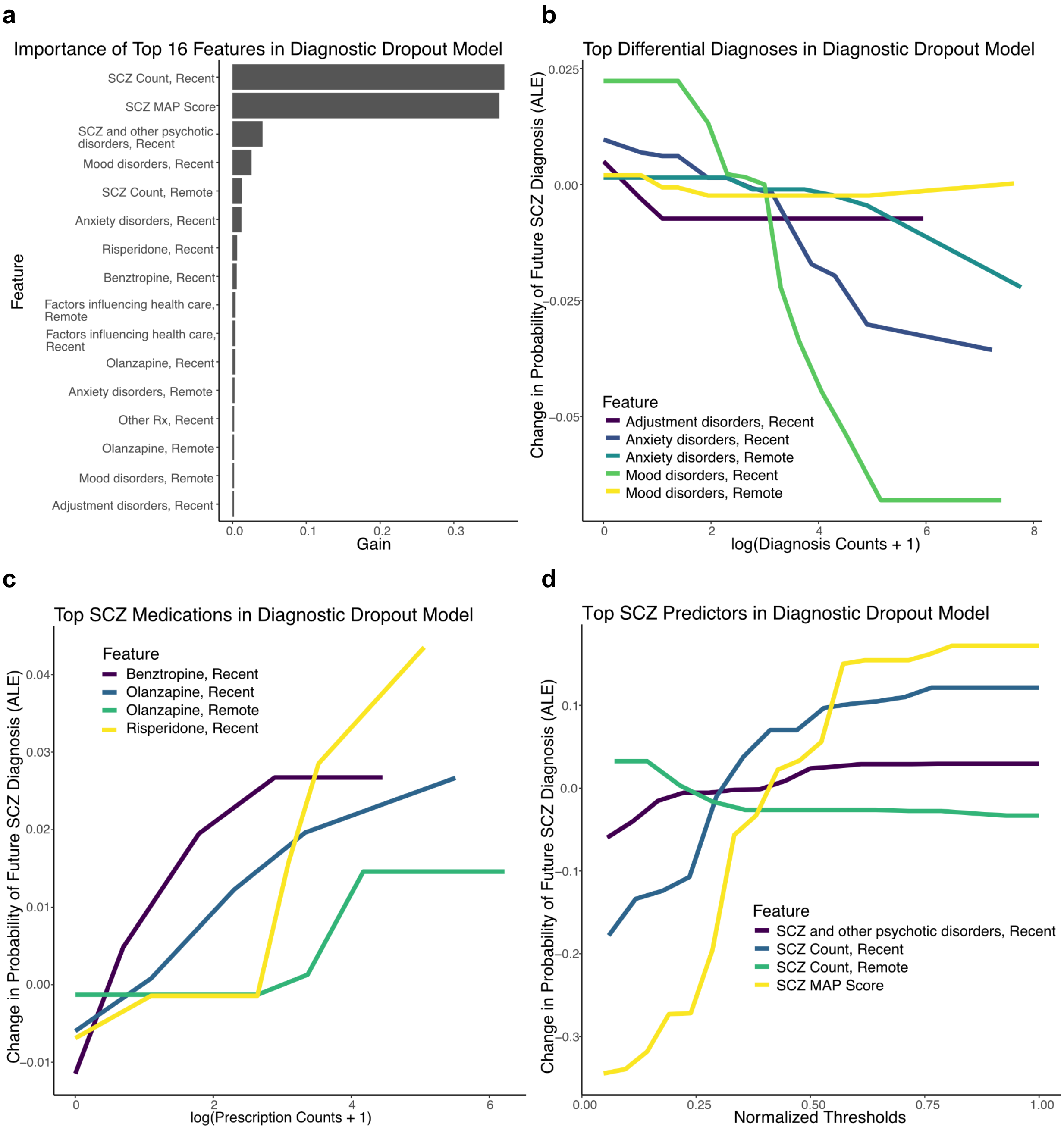


**Figure S2: Interpretation of Model**

**(a)** A bar plot ranking the top predictors by importance in the diagnostic dropout model. The x-axis indicates the gain corresponding to the predictor and the y-axis corresponds to the feature. **(b-d)**. Accumulated local effect plots (ALE) plots corresponding to the top features in the model grouped by theme. The y-axis highlights changes in predictions based on the corresponding predictor. Different colors correspond to different features. Plot **(b)** focuses on log counts of differential diagnoses (x- axis) and their relationship with predicting future schizophrenia codes. Plot **(c)** focuses on log counts of medications (x- axis) and their relationship with predicting future schizophrenia codes. And finally, plot **(d)** focuses on normalized thresholds of predictors corresponding to prior history of schizophrenia diagnoses (x- axis) and their relationship with predicting future schizophrenia codes. Recent and remote refer to data from 5 up to 10 years prior or beyond 10 years from the last encounter, respectively. SCZ Count refers to a count of schizophrenia diagnosis codes as defined per the phecode definition.

### Figure S3

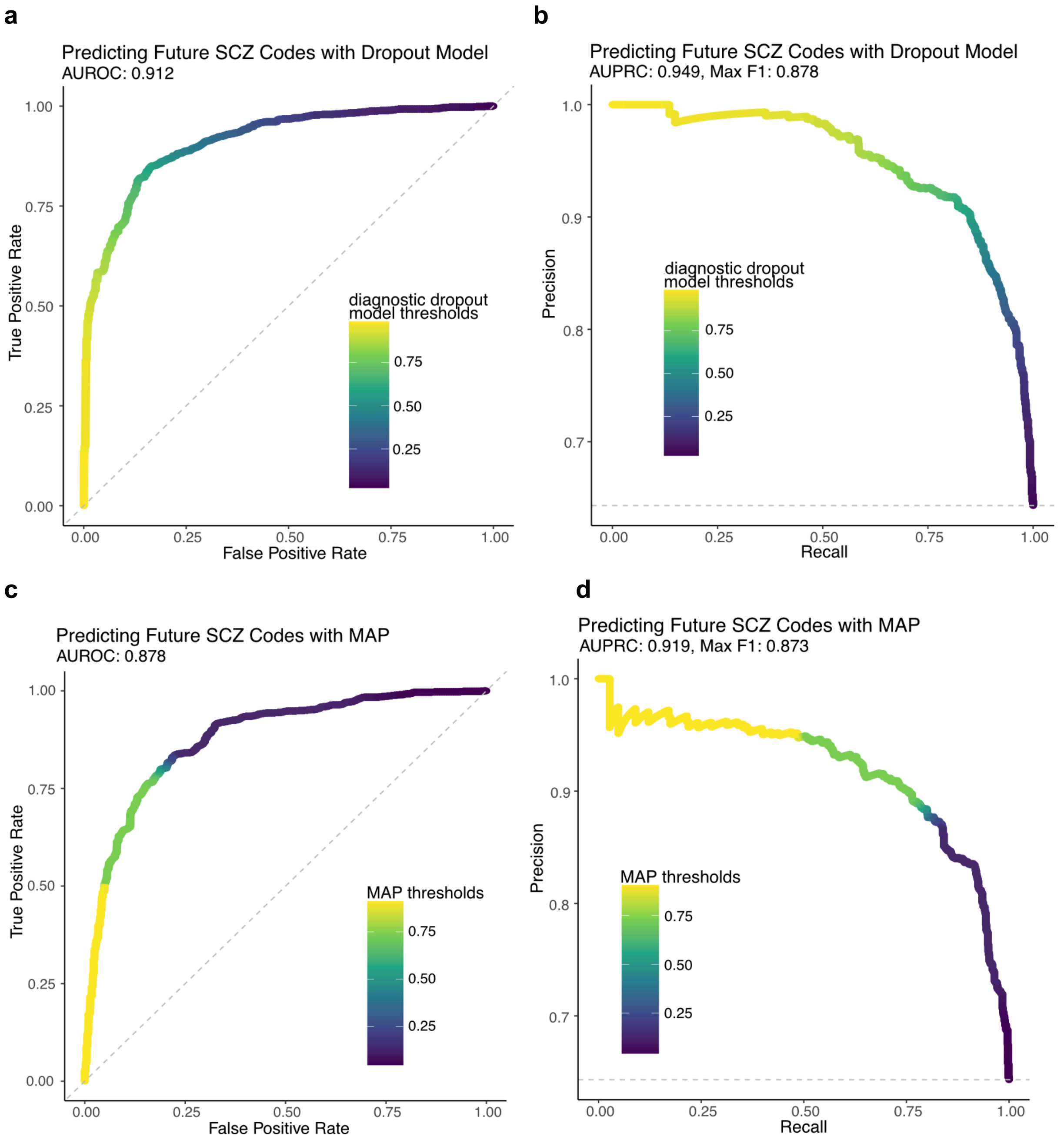


**Figure S3: Performance on Holdout Set**

Receiver operator and precision recall curves assessing the performance of our diagnostic dropout model **(a-b)** and MAP model **(c-d)** on predicting future schizophrenia diagnosis codes. For the precision-recall curves **(b,d)**, the x-axis identifies the recall value, and the y-axis identifies the precision. A dashed line highlights baseline precision of a random classifier. In the receiver operator curves, the x-axis identifies the false positive rate and the y-axis indicates the true-positive rate. The dashed diagonal line corresponds to performance of a random classifier. Points are color coded identifying the threshold that yields the corresponding precision-recall or false positive rate/true positive rate value.

### Figure S4

**a**

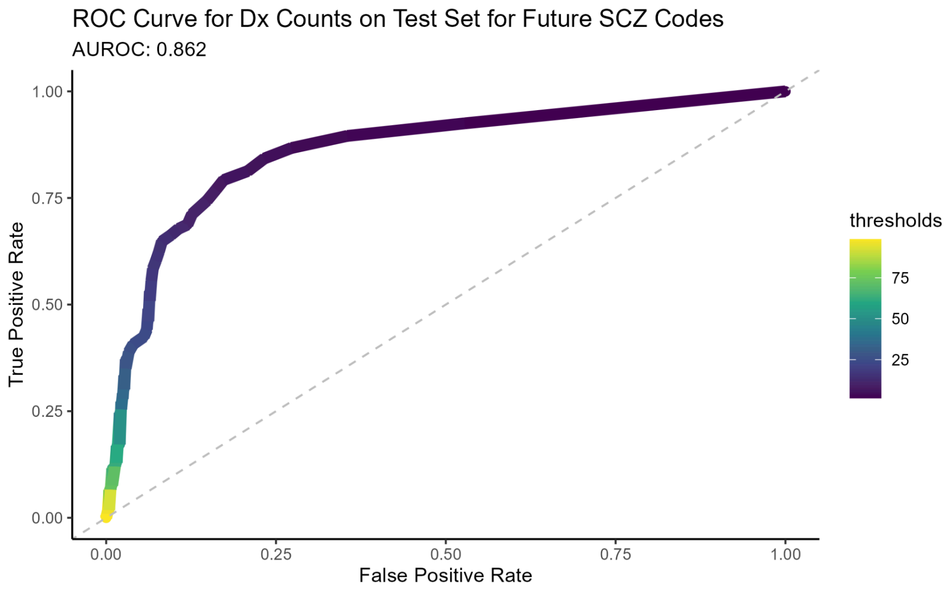


**b**

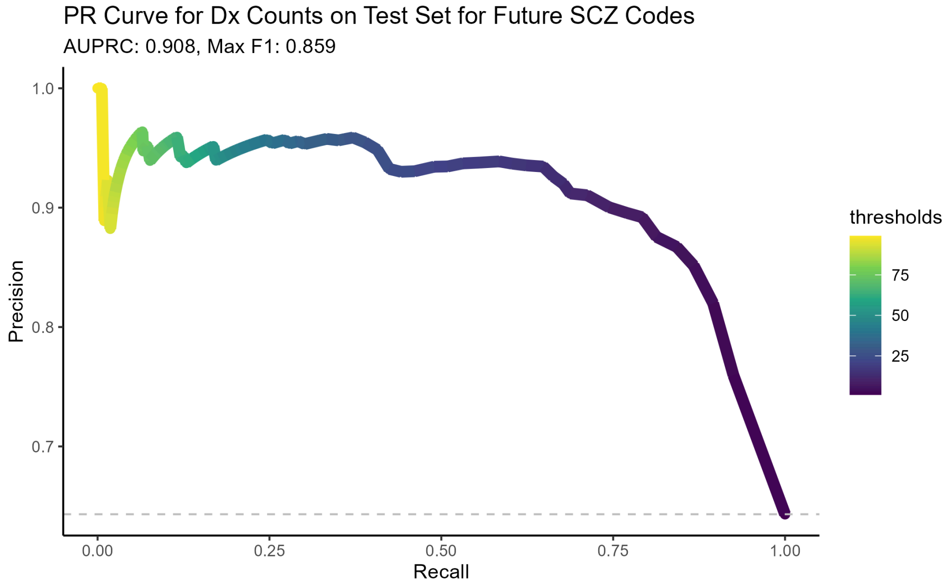


**Figure S4. Performance on Holdout Set.**

Receiver operating characteristic (ROC) **(a)** and precision-recall (PR) curves **(b)** assessing the performance of prior diagnosis code counts for schizophrenia (a-b) on predicting future schizophrenia diagnosis codes. For the PR curve **(b)**, the x-axis identifies the recall value, and the y-axis identifies the precision. A dashed line highlights baseline precision of a random classifier. In the receiver operator curves, the x-axis identifies the false positive rate and the y-axis indicates the true-positive rate. The dashed diagonal line corresponds to performance of a random classifier. Points are color coded identifying the threshold that yields the corresponding precision-recall or false positive rate/true positive rate value.

### Figure S5

**a**

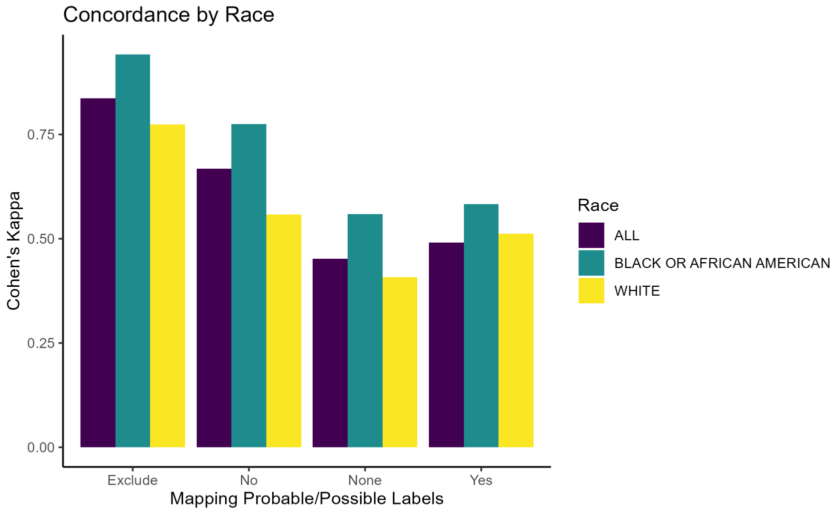


**b**

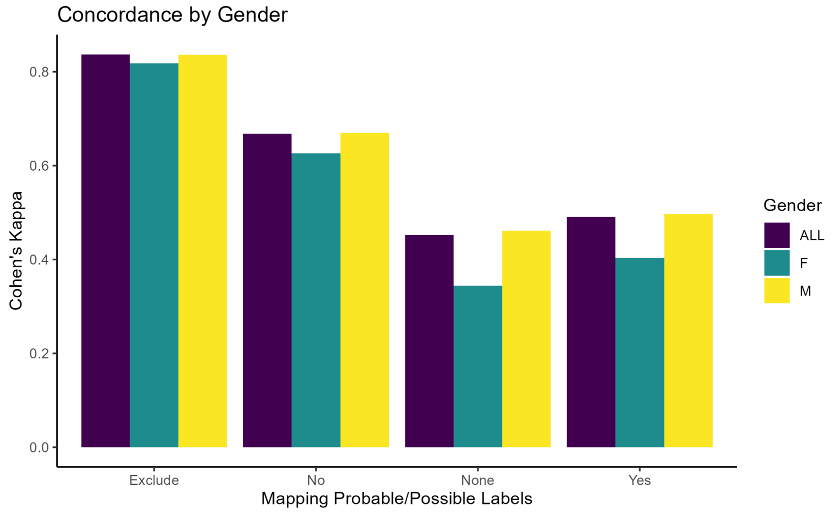


**c**

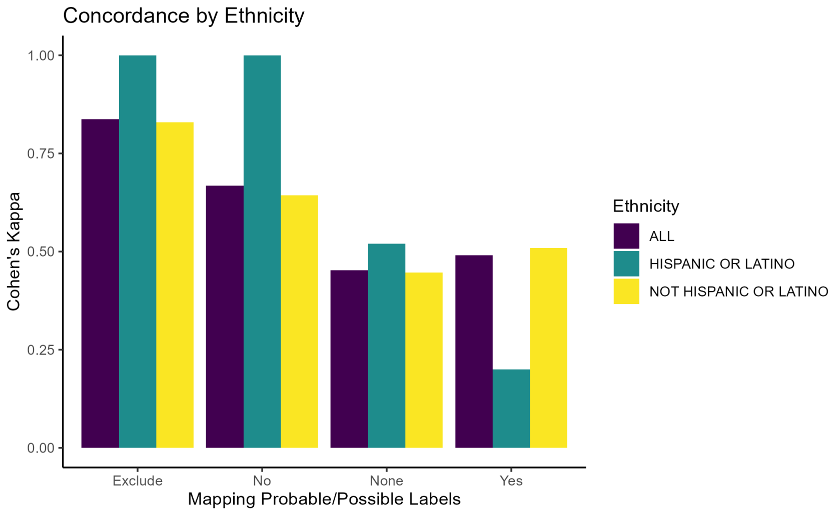


**Figure S5. Clinician concordance was assessed using Cohen's kappa.** First, we estimated overall kappa across all cases ("ALL") and then examined the impact of demographic variables, including race **(a)**, ethnicity **(b)** and gender **(c)**, on agreement. On the x-axis we highlight the labeling system. Reviewers were asked to provide a yes, no, or probable response to the question of whether they think the patient has schizophrenia based on their chart. We compared clinician consensus when classifying schizophrenia by either using the three-category system ("***None***" for Mapping Probable/Possible labels on the x-axis) or simplified two-category systems, where the probable/possible label can be excluded entirely ("***Exclude***") or mapped to yes ("***Yes***") or no ("*No*"). Excluding the probable labels ("***Exclude***") or assigning probable cases to controls ("***No***") exhibit the best interrater kappas across all demographic groups. Note that some groups have very small sample sizes (e.g. Asian individuals) and are not visualized.

### Figure S6

**a**

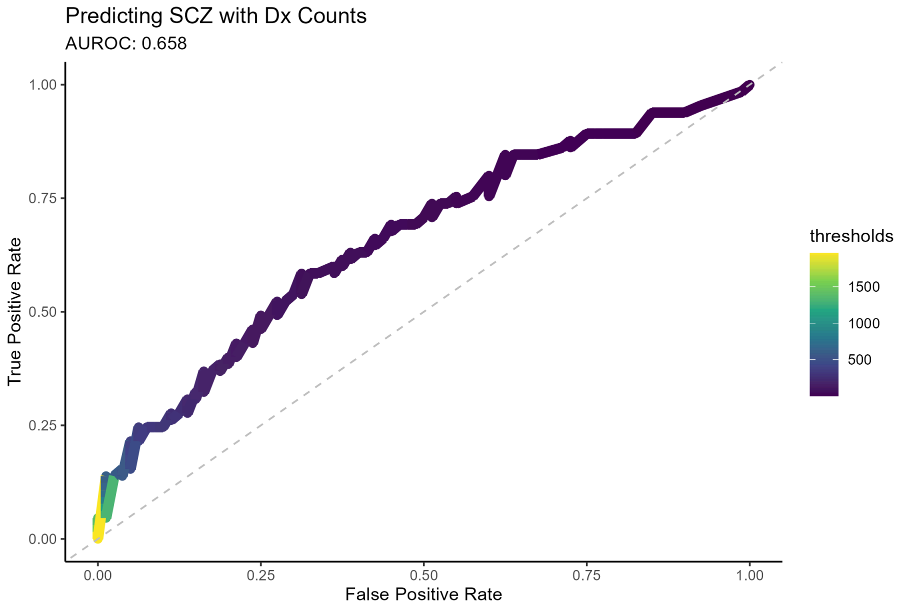


**b**

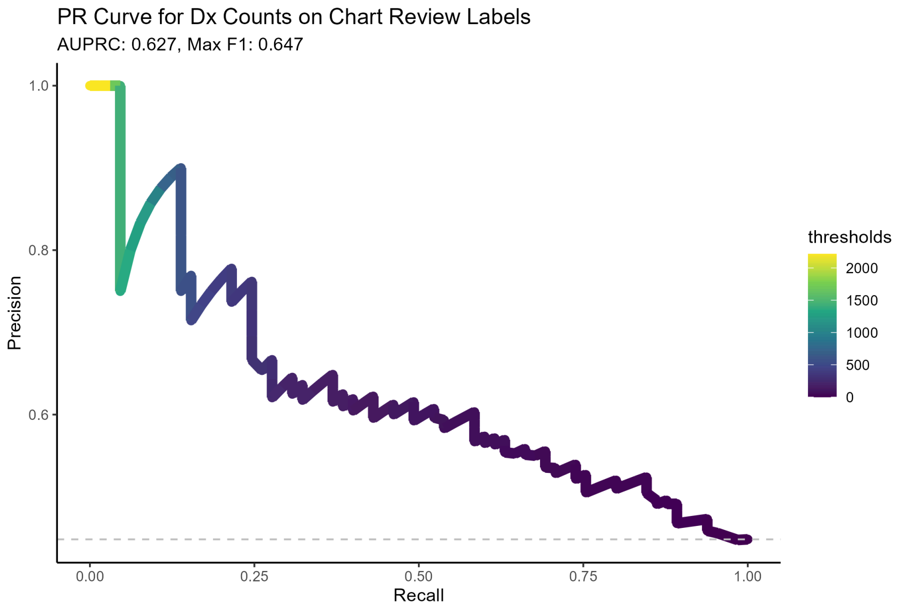


**Figure S6. Performance on Chart Review.**

Receiver operating characteristic (ROC) and precision-recall (PR) curves assessing the performance of using diagnosis code counts **(a,b)** on predicting diagnoses attained through chart review. For the precision-recall curve **(b)**, the x-axis identifies the recall value, and the y-axis identifies the precision. A dashed line highlights baseline precision of a random classifier. In the ROC curves, the x-axis identifies the false positive rate and the y-axis indicates the true-positive rate. The dashed diagonal line corresponds to performance of a random classifier. Points are color coded identifying the threshold that yields the corresponding precision-recall or false positive rate/true positive rate value.

### Figure S7

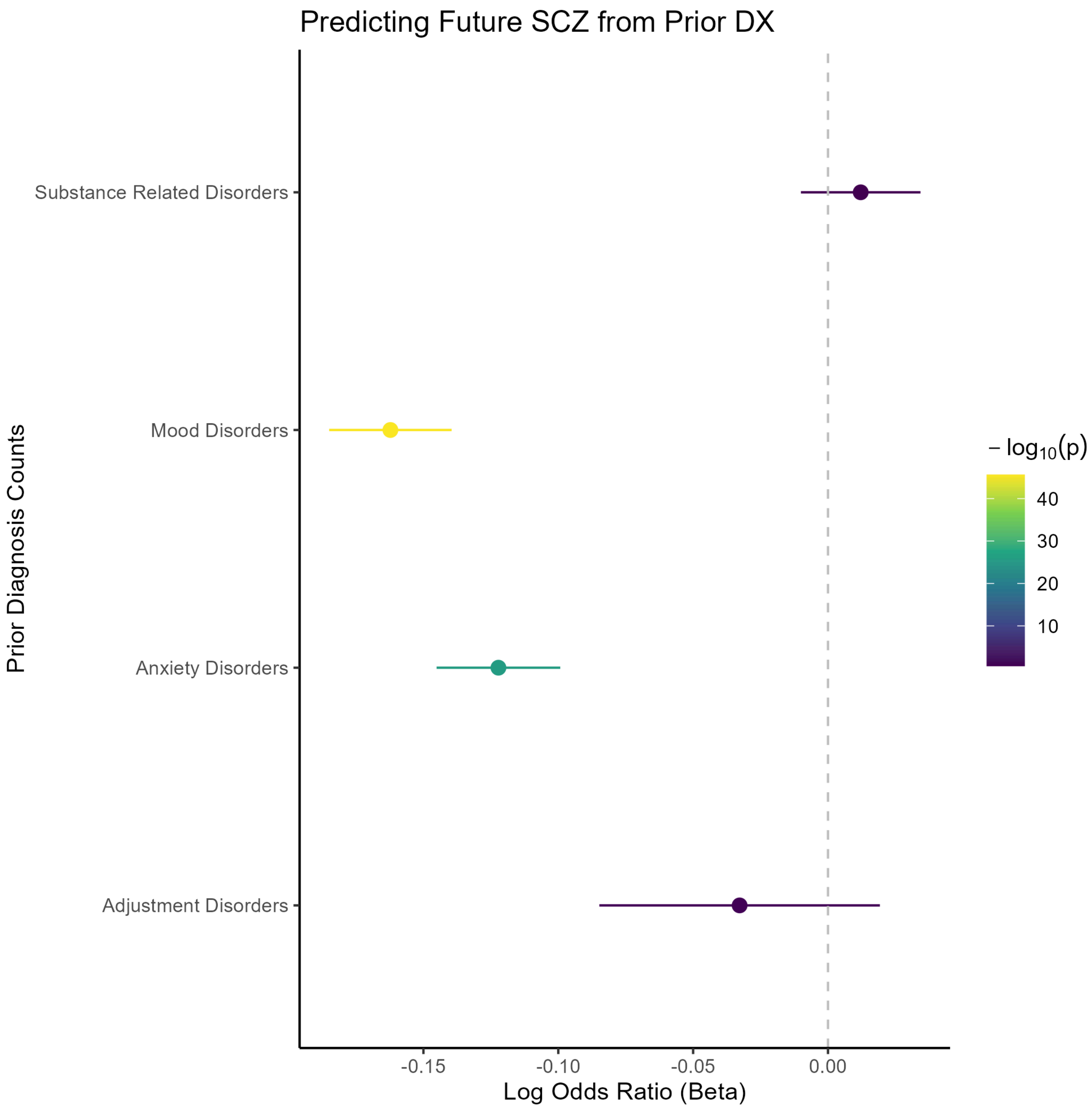


**Figure S7. Auxiliary Logistic Regression Model Predicting Future Schizophrenia Codes.**
Forest plot for differential diagnoses for predicting future schizophrenia diagnosis codes. The dashed gray vertical line corresponds to 0; 95% confidence intervals are plotted for adjustment disorders, anxiety disorders, mood disorders and substance related disorders. Diagnosis counts are computed over a five year period and log transformed. The color of the confidence interval corresponds to the p-value on the logarithmic scale. The x-axis denotes the log odds ratios in the logistic regression model.

Figure S8

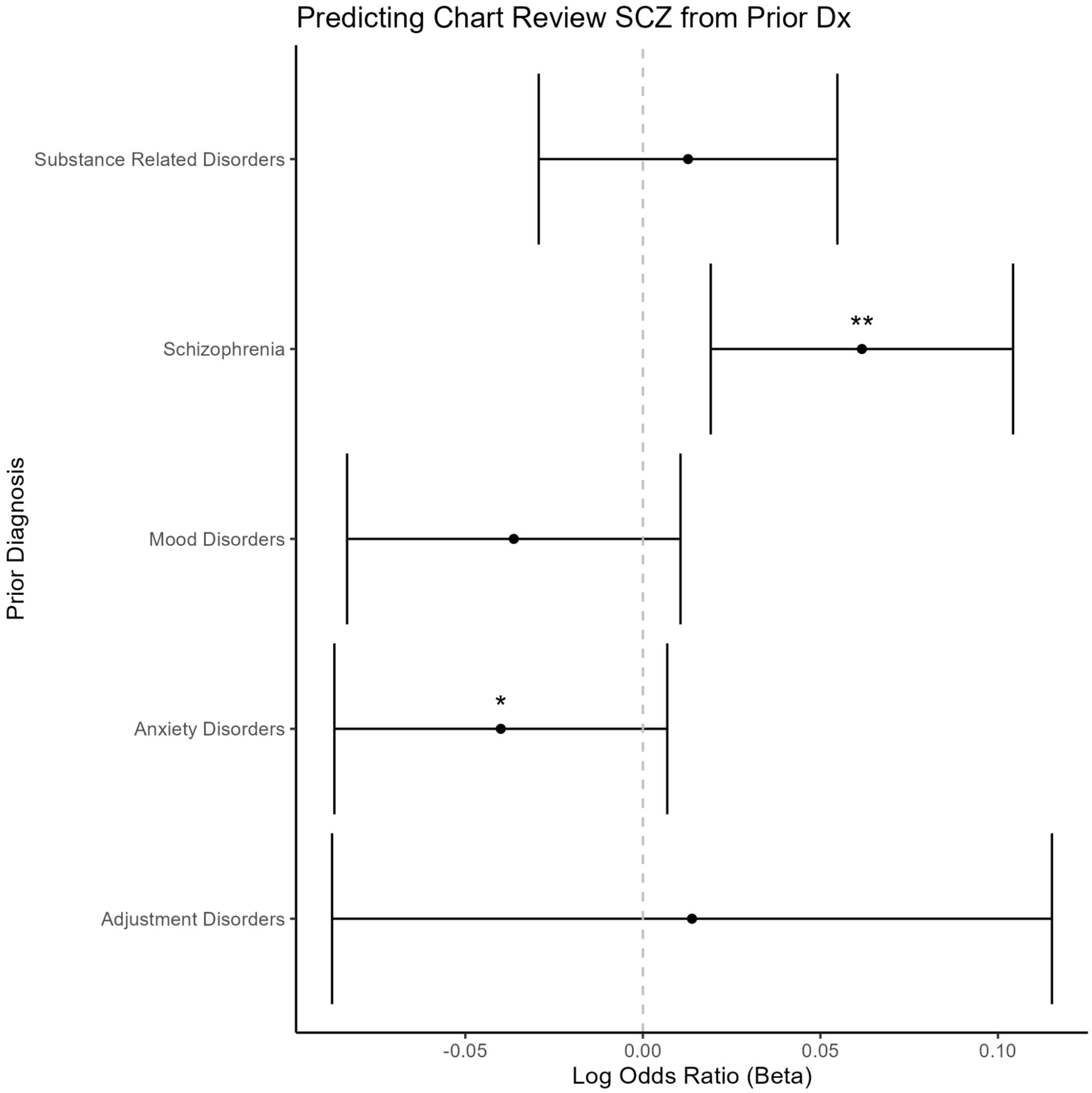


**Figure S8. Logistic Regression Model Predicting Chart Review Labels.**
Forest plot for differential diagnoses for predicting chart review labels. The dashed gray vertical line corresponds to 0 and 95% confidence intervals are plotted for adjustment disorders, anxiety disorders, mood disorders, schizophrenia and substance related disorders. Diagnosis counts are computed over a five year period and log transformed. The x-axis denotes the log odds ratios in the logistic regression model. Statistical tests are one-sided with the direction determined by the diagnosis dropout model. (* $p < .05$, ** $p < .01$, *** $p < .001$)

Figure S9

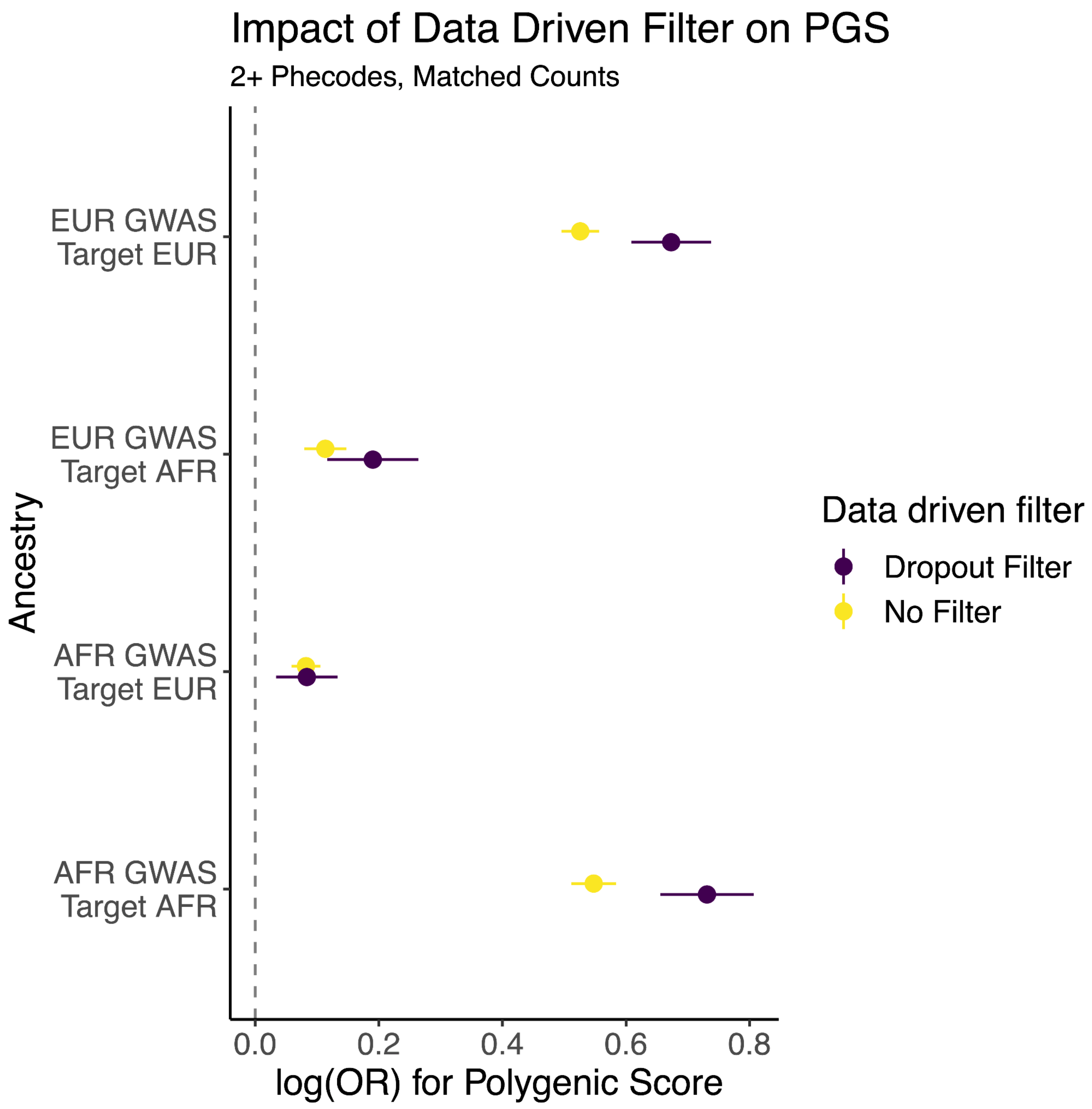


**Figure S9. Improvement of PGS Performance when applying Data-Driven Filter to PheCode Counts.** Forest plot for the differences in effect size of polygenic score predictor between applying and not applying our data driven filter based on the diagnostic dropout model for schizophrenia cases defined through phecode (grouped diagnosis code) counts. The x-axis indicates the difference in the log odds ratio of the polygenic score. The y-axis corresponds to the training cohort GWAS for constructing the PGS as well as the validation cohort. The color indicates whether a filter was used. While we plot two sided confidence intervals, statistical tests are one-sided. A dashed vertical line corresponds to the null hypothesis of no difference in effect between filters.

Figure S10

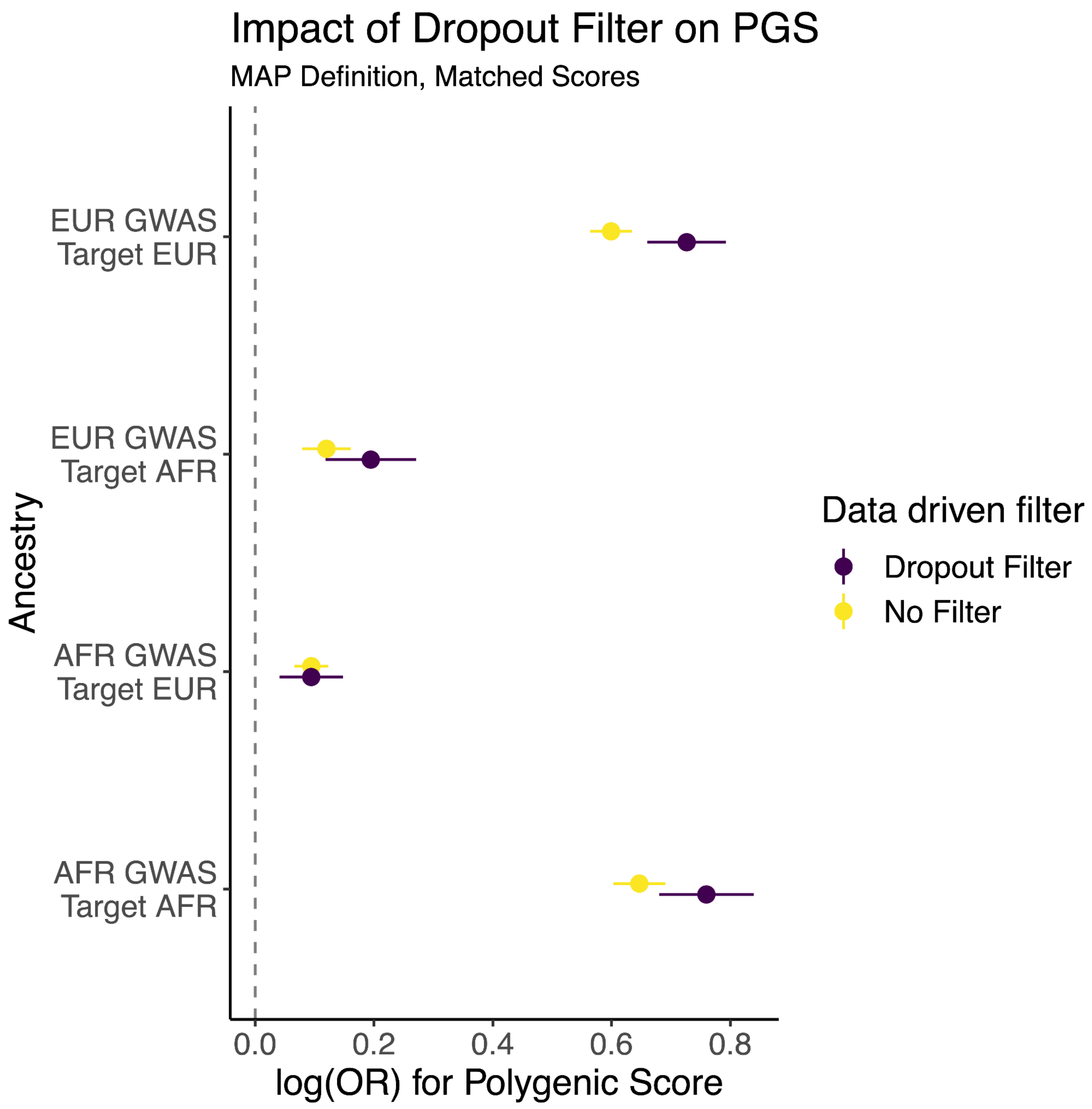


**Figure S10. Improvement of PGS Performance when applying Data-Driven Filter to MAP.** Forest plot for the differences in effect size of polygenic score predictor between applying and not applying our data driven filter based on the diagnostic dropout model for schizophrenia cases defined through MAP. The x-axis indicates the difference in the log odds ratio of the polygenic risk score. The y-axis corresponds to the training cohort GWAS for constructing the PRS as well as the validation cohort. The color indicates whether a filter was used. While we plot two sided confidence intervals, statistical tests are one-sided. A dashed vertical line corresponds to the null hypothesis of no difference in effect between filters.

Figure S11

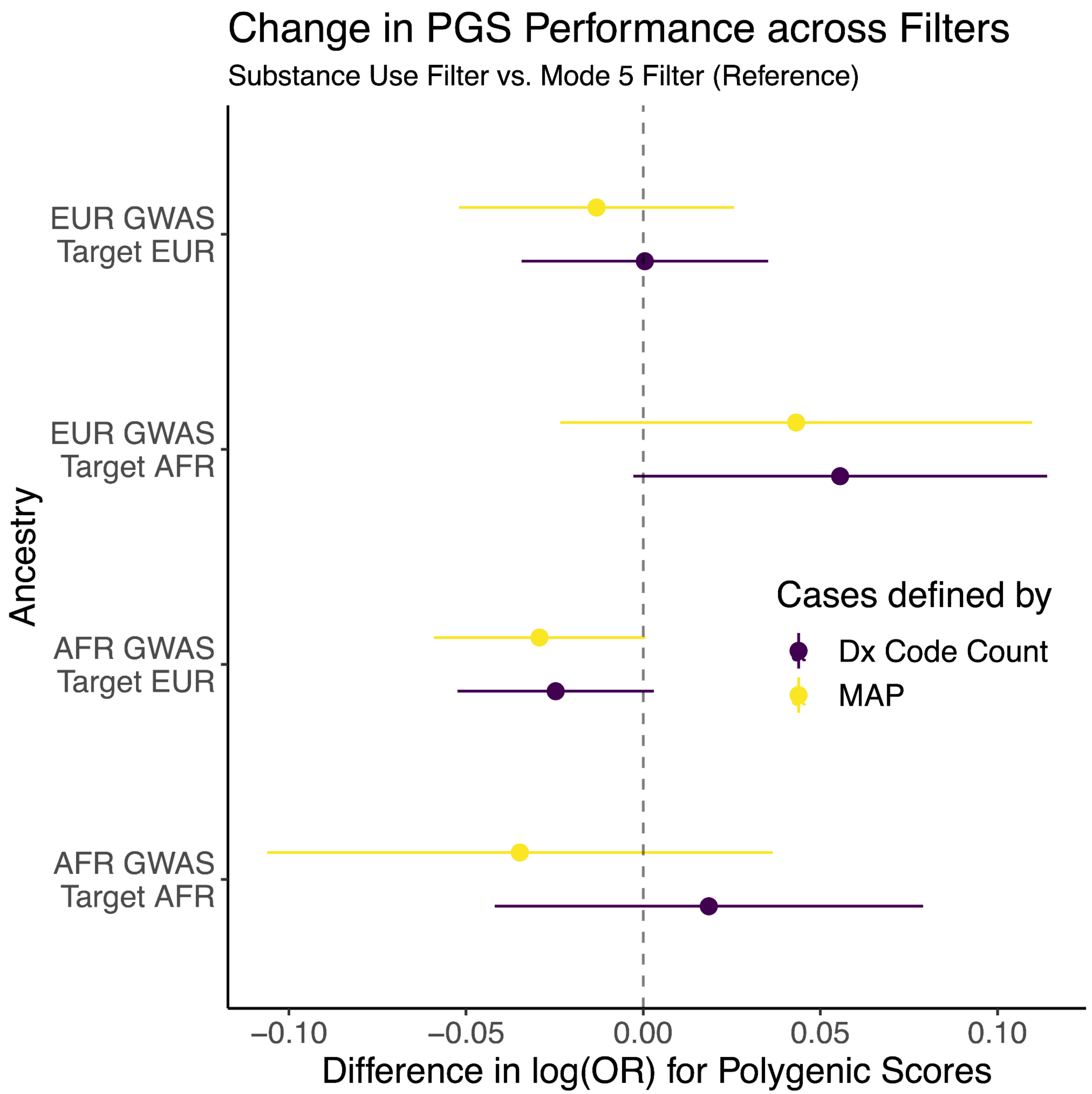


**Figure S11. Comparing Addition of Substance Use Filter to Mode 5.**
Forest plot for the differences in effect size of polygenic risk score predictor between applying different filters for schizophrenia cases. We compare the addition of a substance use filter on top of a Mode 5 filter, removing cases where bipolar disorder diagnoses are more commonly observed than schizophrenia diagnoses across the five most recent bipolar/schizophrenia diagnoses. The x-axis indicates the difference in the log odds ratio of the polygenic score. The y-axis corresponds to the training cohort GWAS for constructing the PGS as well as the validation cohort. The color of the confidence interval corresponds to whether the MAP or diagnosis code count definition was used for identifying schizophrenia cases. A dashed vertical line corresponds to the null hypothesis of no difference in effect between filters. P-values are annotated as follows: * p < .05, ** p < .01, *** p < .001.

Figure S12

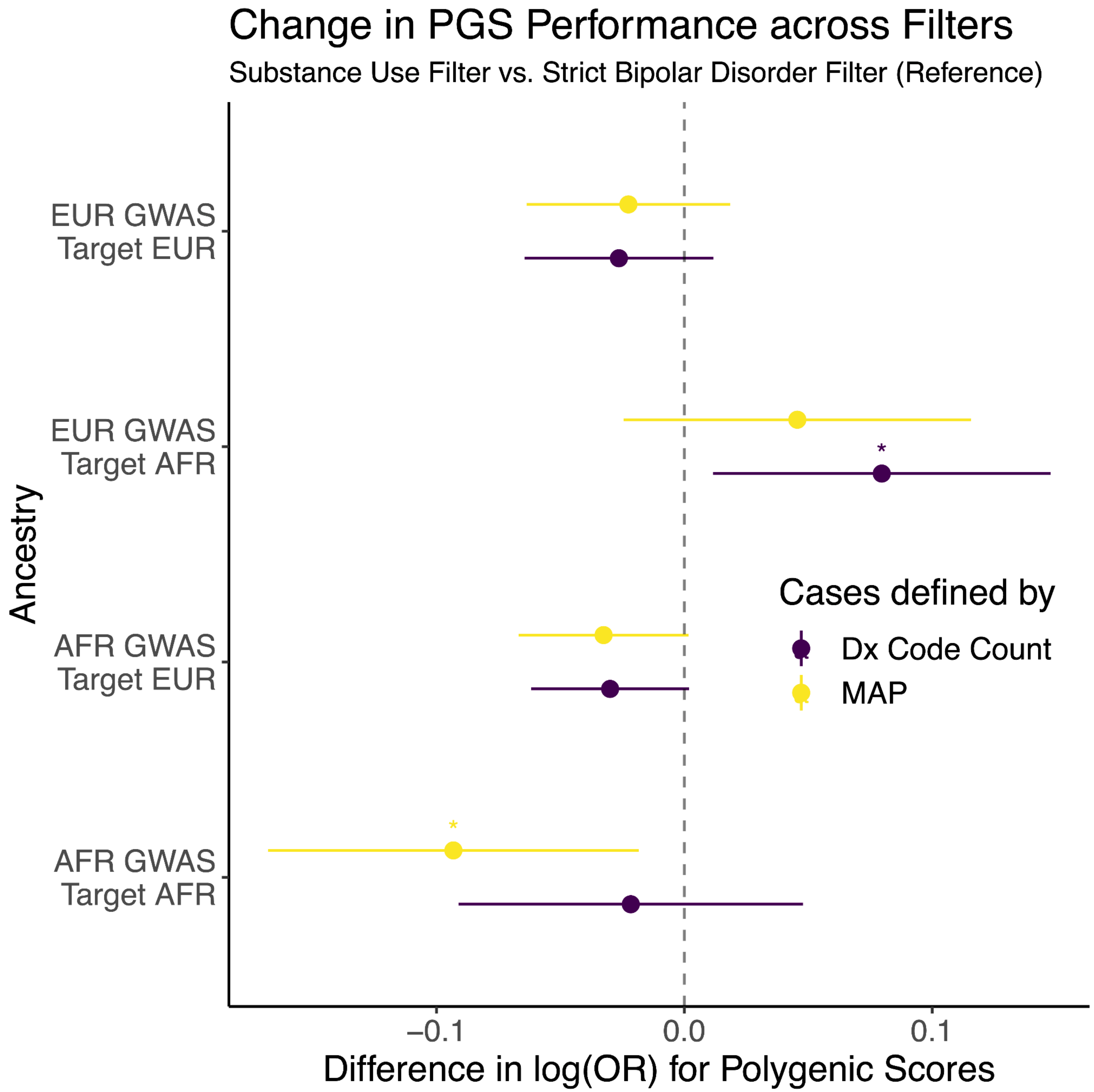


**Figure S12. Comparing Addition of Substance Use Filter to Strict Bipolar Disorder Filter.**
Forest plot for the differences in effect size of polygenic risk score predictor between applying and different filters for schizophrenia cases. We compare the addition of a substance use filter on top of a strict bipolar disorder filter, removing all cases if a bipolar disorder diagnosis appears in the chart. The x-axis indicates the difference in the log odds ratio of the polygenic risk score. The y-axis corresponds to the training cohort GWAS for constructing the PGS as well as the validation cohort. The color of the confidence interval corresponds to whether the MAP or diagnosis code count definition was used for identifying schizophrenia cases. A dashed vertical line corresponds to the null hypothesis of no difference in effect between filters. P-values are annotated as follows: * p < .05, ** p < .01, *** p < .001.

### eMethods

#### CSP 572 and MVP

Only schizophrenia cases that were exclusively enrolled in CSP #572 and not the MVP were retained in the analysis. To account for sample overlap in controls between CSP #572 and the MVP, we fit a Gaussian mixture model to generate probabilities that an individual was a control in both cohorts based on their polygenic score, and performed importance sampling to account for any bias in sample overlap between the two groups

#### Predicting Diagnostic Dropout

For details on the schematic of the model, we refer the reader to **Figure 1**. Predictors in the model included MAP scores for schizophrenia (not visualized in **Figure 1** because MAP scores are not required for modeling diagnostic dropout for in other cohorts), prior diagnosis and prescription code counts, where the latter two categories of features were binned temporally to separate newer and older codes that appear in the electronic health record from 5 to 10 years prior to the most recent outpatient encounter and from more than 10 years prior to the most recent outpatient encounter. MAP scores were computed by the MVP Data Core Team, where we utilized the most recent release of MAP available, (v19.2), which excludes updates to patient chart data past October 2019. We fine tuned parameters through cross-validation. Hyperparameter selection was determined by 5-fold cross-validation on the training set. Hyperparameters included the number of rounds (100 or 200), gamma (0 or .5), eta (.1 or .3), max depth (2 or 3), column sample by tree (.7 or .9), subsample (.6 or .8). The minimum child weight equaled 1. We ran the train function from the caret package in R[1], with the 'xgbTree' method as a classification problem, where the model predicts if 2 or more schizophrenia diagnosis codes appear in the last 5 years of a patient's chart. We assessed model performance on a 10% holdout set and compared the performance of our approach to other methodologies, including MAP[2] and diagnosis code counts[3].

Predictors included diagnosis codes from outpatient encounters, aggregated through Clinical Classification Software Revised (CCS) codes[4]. Schizophrenia phecodes were also used as predictors in the model[5]. Outpatient medications were aggregated through the ingredient level using a table provided by the MVP Core Data Team. In addition, we also leveraged the phecode definition for schizophrenia in the model as well. Although MAP scores are not strictly required for the diagnostic dropout model, MAP scores were integrated into the diagnostic dropout model under the hypothesis that their inclusion would help improve performance of our model to outperform MAP and other benchmarks. Patients with more than 100 schizophrenia codes over the 5 year period highlighted in green or yellow in **Figure 1** were excluded from the training of the model.

To complement our XGBoost analysis, we also trained a logistic regression model, predicting the presence of 2 or more future schizophrenia diagnosis codes over the five most recent years of a patient's chart, leveraging prior diagnosis code counts over a five year period for schizophrenia, substance use disorders, anxiety disorders, mood disorders and adjustment disorders. All diagnosis groups were log transformed in the model.

#### Chart Review Validation

To select charts for review, we leveraged MAP scores for schizophrenia, classifying cases into three probability levels: low (if score < 25%), medium (if score between 25% and 75%), or high (if score >75%) likelihood of having SCZ diagnosis. We then defined a subset of 146 charts from age-, race-, and gender-matched individuals with an approximately uniform distribution of binned MAP scores, (low SCZ scores, (0,25%), n = 49;

moderate SCZ scores, (25%,75%), n = 43; high SCZ scores, (75%,100%), n = 54. For each chart, clinicians were asked to determine diagnoses **(Supplementary Table S3**) based on review of available demographic information (age, gender, race, ethnicity), ICD codes for both medical and psychiatric diagnoses (ICD, ICD date), medication history (drug name, drug dose, prescription/administration date), available labs (test name, test date, lab value, lab reference value), and clinicians' notes from inpatient and outpatient encounters. Inter-rater reliability was estimated using Cohen's kappa.

For chart review analyses, we defined schizophrenia cases as patients who received two certain diagnoses of schizophrenia through chart review. To assess statistical significance of our diagnostic dropout model, we also trained a logistic regression model on our validation set to predict if patients have two "Yes schizophrenia" ratings, using log diagnosis code counts, log odds ratios of the diagnosis dropout scores and log odds ratios of the MAP scores.

#### Genotyping, quality control and imputation

MVP Genotyping, initial quality control, and imputation is handled by the MVP Data Team as previously described[6–8]. For completeness, we provide a brief overview of the methodologies employed. This study leveraged release 4 of the MVP, which contains genotyping data from blood samples for more than 650,000 individuals. The MVP 1.0 custom Genotyping array was used for genotyping, where the APT software 2.11.3 was used for genotype calling. Genotyping array. Prephasing was performed using EAGLE v2[9] Genotype quality control metrics encompass: sex check, rare heterozygous adjustment, plate normalization and sample labeling. The MVP Data Team performed ancestry-specific principal component analysis using PLINK 2.0[10] and computed relatedness (kinship coefficient) with KING 2.1[11]. Ancestry was computed using HARE[12] (genetically inferred ancestry leveraging the top 30 principal components and ADMIXTURE[13]) and SHAPEIT4[14] version 4.1.3 was used to phase genotypes. Principal component analysis to generate ancestral principal components was performed using EIGENSOFT v.6[15,16].Pre-imputation QC filtered out variants that had significant missingness (> 20%), were monomorphic, or had substantial variation in frequency across batches. The MVP Data Team imputed genotypes with either the Trans-Omics for Precision Medicine (TOPMed) reference panel[17] or the 1000 Genomes Project phase 3, version 5 reference panel[18]. We then performed follow-up quality control filters where we only included samples with sample call rates exceeding 98.5%, required that that the sample heterozygosity rates deviate at most four standard deviations from the samples' heterozygosity rate mean and filtered out related samples/samples with cryptic relationship by leveraging a cut-off value of .0884 for the kinship coefficient (first we removed samples with multiple relationships; subsequently, we filtered out samples with the highest missingness rates from the remaining relationship pairs). We performed variant-level filtering retaining SNPs such that the MAF exceeded 0.005, the effective minor allele count was at least 30, the variant call rates exceeded 95%, the Hardy-Weinberg equilibrium test p-value exceeded $5 \times 10^{-8}$, and the imputation $R^2$ exceeded 0.4. We leveraged logistic regression for performing the GWAS analysis with PLINK 2.0. We performed separate analyses on different genetic ancestries and adjusted for confounders, including sex, age and the top 10 principal components.

#### Assessment of Data-driven Phenotypes with Polygenic Scores

Based on our model, we defined the dropout filter using recent diagnoses, as remote diagnoses contributed less to diagnostic dropout than recent diagnoses. With the exception of the mode 5 filter, substance use and bipolar disorder filters were implemented such that the presence of any such diagnosis would result in the exclusion of that case.

When assessing filter performance, since filtered schizophrenia cases could have a different MAP scores/diagnosis code count distribution than non-filtered cases, we applied importance sampling to create comparable MAP score/diagnosis code distributions between filtered and unfiltered schizophrenia cases. More specifically, MAP score/diagnosis code distributions for potential schizophrenia cases that passed and failed the filter were estimated through Gaussian kernel density estimation. An importance weight for each case that failed the filter was derived by taking the ratio of the probability density from the group that passed the filter divided by the probability density from the group that failed the filter, where each function was evaluated based on the MAP or diagnosis code count for the case of interest. Importance weights were then normalized to sum to 1. Cases that passed the filter and controls had uniform weights (with the possible exception for African ancestry controls, where weights were derived as described above to account for potential sample overlap). Bootstrap samples were then generated, where samples were selected with replacement with the corresponding importance weight and the resulting simulated dataframe matched the same number of rows as the original dataframe. Confidence intervals were constructed by performing 2,000 bootstrap simulations.

When comparing the impact of a filter, we used a paired test, by comparing the effect of the filter when the controls are identical. To complement our analysis comparing the performance of the polygenic scores when utilizing and not utilizing the filter, we also compare our data-driven filter with other filtering approaches referenced in the literature[19] to filter out any bipolar disorder patients or substance use disorder patients in our analysis[19,20], as patients with either of these diagnoses can potentially be misdiagnosed with schizophrenia.

#### Measuring Dilution with PheMED

Confidence intervals for measuring dilution through PheMED were generated by using 2,000 bootstrap simulations. For PheMED analyses, we created two non-overlapping GWAS's for European and African ancestries where for one analysis, cases passed the diagnosis dropout filter and for the other analysis, cases failed the diagnosis dropout filter. Schizophrenia cases were defined according to the standard MAP threshold, and otherwise excluded from the analysis. To perform a cross-ancestry meta-analysis of the dilution estimates in a joint PheMED analysis, we computed a weighted average of the dilution values from not applying the diagnostic dropout filter for African and European ancestry GWAS's, where the weights were derived based on an extension of the invariance weights meta-analysis that accommodates correlated estimates[21]. P values were then computed using the methodology described in the PheMED paper[7].

### Supplementary Results

#### Top Medications in Model

Here, we extend our discussion on the top medications found in our diagnostic dropout model. In addition to olanzapine, our model also utilizes prescription counts for risperidone, an effective and versatile alternative often preferred during inpatient admissions for rapid symptom stabilization with good long-acting injectable formulation follow-up options. Both olanzapine and risperidone have been well-established in the treatment of schizophrenia[22]. Additionally, benztropine, a medication typically prescribed to manage side effects associated with long-term use of high-dose first-generation antipsychotics[23], is also identified as a positive predictor for the later appearance of schizophrenia diagnosis codes in the patient's chart. In contrast, first-generation antipsychotics (like haloperidol, fluphenazine and perphenazine) were not identified by our model. This may be due to a lower prevalence of first-generation antipsychotic prescriptions in the dataset, possibly reflecting a shift in prescribing patterns over time. As these medications are typically associated with more severe extrapyramidal side effects[24], it is likely that patients have been transitioned to second-generation antipsychotics, which offer a more favorable side effect profile, especially in the more recent years of observation.

#### Rater Agreement

Cohen's interrater kappa (κ) was initially calculated at 0.452 when reviewers classified schizophrenia into three categories: yes, no, or possible, as we did not require chart reviewers to provide a diagnosis if they were uncertain. This kappa value indicates moderate agreement among reviewers, as there was some variation when including the "possible" category. Additionally, concordance was consistent across cases and was not influenced by factors such as ethnicity, race, or gender, indicating that the reviewers' classifications were unbiased with respect to these demographic variables (**Supplementary Figure S5, eMethods**) and did not introduce biases beyond what existed *a priori* in the patients' charts.

#### Selecting Top Differential Diagnoses

We also note that our model identifies adjustment disorders as contributing to the risk of diagnostic dropout for schizophrenia. This finding has been reported in a prior study[25], but we chose not to include adjustment disorders as part of the data-driven filter as the importance of this feature was much smaller than mood and anxiety disorders and did not achieve statistical significance in our auxiliary logistic regression model for predicting the presence of 2 or more schizophrenia diagnosis codes in the 5 most recent years of the patient's chart (**Methods)**.

We stress that data-driven filters for defining robust phenotypes could vary across different electronic health record datasets and this can pose challenges in multi-site collaborative projects. For example, in a prior work that predicted diagnoses that appear subsequent to a patient's schizophrenia diagnosis, anxiety disorders did not appear in the model[25], even though they have been well established as a differential diagnosis for schizophrenia[26]. Here, we present a data-driven methodology that can identify diagnoses that directly relate to diagnostic uncertainty, as overly restrictive definitions can unnecessarily decrease the sample size in our analyses **(Supplementary Table S1)**.

### Supplementary Acknowledgements


VA Million Veteran Program:
Core Acknowledgements for Publications
May 2024

MVP Program Office

- Sumitra Muralidhar, Ph.D., Program Director

  US Department of Veterans Affairs, 810 Vermont Avenue NW, Washington, DC 20420

- Jennifer Moser, Ph.D., Associate Director, Scientific Programs

  US Department of Veterans Affairs, 810 Vermont Avenue NW, Washington, DC 20420

- Jennifer E. Deen, B.S., Associate Director, Cohort & Public Relations

  US Department of Veterans Affairs, 810 Vermont Avenue NW, Washington, DC 20420

MVP Executive Committee

- Co-Chair: Philip S. Tsao, Ph.D.
  VA Palo Alto Health Care System, 3801 Miranda Avenue, Palo Alto, CA 94304
- Co-Chair: Sumitra Muralidhar, Ph.D.
  US Department of Veterans Affairs, 810 Vermont Avenue NW, Washington, DC 20420
- J. Michael Gaziano, M.D., M.P.H.
  VA Boston Healthcare System, 150 S. Huntington Avenue, Boston, MA 02130
- Elizabeth Hauser, Ph.D.
  Durham VA Medical Center, 508 Fulton Street, Durham, NC 27705
- Amy Kilbourne, Ph.D., M.P.H.
  VA HSR&D, 2215 Fuller Road, Ann Arbor, MI 48105
- Michael Matheny, M.D., M.S., M.P.H.
  VA Tennessee Valley Healthcare System, 1310 24th Ave. South, Nashville, TN 37212
- Dave Oslin, M.D.
  Philadelphia VA Medical Center, 3900 Woodland Avenue, Philadelphia, PA 19104
- Deepak Voora, MD
  Durham VA Medical Center, 508 Fulton Street, Durham, NC 27705

MVP Co-Principal Investigators

- J. Michael Gaziano, M.D., M.P.H.

  VA Boston Healthcare System, 150 S. Huntington Avenue, Boston, MA 02130

- Philip S. Tsao, Ph.D.

  VA Palo Alto Health Care System, 3801 Miranda Avenue, Palo Alto, CA 94304

MVP Core Operations

- Jessica V. Brewer, M.P.H., Director, MVP Cohort Operations

  VA Boston Healthcare System, 150 S. Huntington Avenue, Boston, MA 02130

- Mary T. Brophy M.D., M.P.H., Director, VA Central Biorepository

  VA Boston Healthcare System, 150 S. Huntington Avenue, Boston, MA 02130

- Kelly Cho, M.P.H, Ph.D., Director, MVP Phenomics

  VA Boston Healthcare System, 150 S. Huntington Avenue, Boston, MA 02130

- Lori Churby, B.S., Director, MVP Regulatory Affairs

  VA Palo Alto Health Care System, 3801 Miranda Avenue, Palo Alto, CA 94304

- Scott L. DuVall, Ph.D., Director, VA Informatics and Computing Infrastructure (VINCI)

  VA Salt Lake City Health Care System, 500 Foothill Drive, Salt Lake City, UT 84148

- Saiju Pyarajan Ph.D., Director, Data and Computational Sciences

  VA Boston Healthcare System, 150 S. Huntington Avenue, Boston, MA 02130

- Robert Ringer, Pharm.D., Director, VA Albuquerque Central Biorepository

  New Mexico VA Health Care System, 1501 San Pedro Drive SE, Albuquerque, NM 87108

- Luis E. Selva, Ph.D., Director, MVP Biorepository Coordination

  VA Boston Healthcare System, 150 S. Huntington Avenue, Boston, MA 02130

- Shahpoor (Alex) Shayan, M.S., Director, MVP PRE Informatics

  VA Boston Healthcare System, 150 S. Huntington Avenue, Boston, MA 02130

- Brady Stephens, M.S., Principal Investigator, MVP Information Center

  Canandaigua VA Medical Center, 400 Fort Hill Avenue, Canandaigua, NY 14424

- Stacey B. Whitbourne, Ph.D., Director, MVP Cohort Development and Management

  VA Boston Healthcare System, 150 S. Huntington Avenue, Boston, MA 02130

MVP Publications and Presentations Committee

- Co-Chair: Themistocles L. Assimes, M.D., Ph. D

  VA Palo Alto Health Care System, 3801 Miranda Avenue, Palo Alto, CA 94304

- Co-Chair: Adriana Hung, M.D.; M.P.H

  VA Tennessee Valley Healthcare System, 1310 24th Ave. South, Nashville, TN 37212

- Co-Chair: Henry Kranzler, M.D.

  Philadelphia VA Medical Center, 3900 Woodland Avenue, Philadelphia, PA 19104

19. Bigdeli TB, Voloudakis G, Barr PB, et al. Penetrance and pleiotropy of polygenic risk scores for schizophrenia, bipolar disorder, and depression among adults in the US veterans affairs health care

system. *JAMA Psychiatry*. 2022;79(11):1092.
